# Structural variant discovery and diagnostic impact in rare diseases from short-read and long-read sequencing

**DOI:** 10.64898/2026.06.22.26356238

**Authors:** Alba Sanchis-Juan, Yulia Mostovoy, Sarah L Stenton, Vijay S Ganesh, Ben Weisburd, Alex Yenkin, Nehir E Kurtas, Xuefang Zhao, Eren Shin, Philip M Boone, Hang Su, Arthur S Lee, Rachita Yadav, Kirsten Allan, Emanuela Argilli, Christina Austin-Tse, Brenda J Barry, Samantha Baxter, Alan H Beggs, Katrina M Bell, Benjamin Blankenmeister, Carsten G Bönnemann, Catherine A Brownstein, Kinga M Bujakowska, Elizabeth Carbonell, Sandra T Cooper, Laura E Covill, Stephanie DiTroia, Sandra Donkervoort, Elizabeth C Engle, Lyndon Gallacher, Casie A Genetti, Joseph G Gleeson, Bin Guan, Stacey Hall, Friedhelm Hildebrandt, Robert B Hufnagel, Julie A Jurgens, Akanksha Khorgade, Gabrielle Lemire, Emily Liau, Jialan Ma, Jill A Madden, Brian Mangilog, Brandy M McNulty, Olfa Messaoud, Shloka Negi, Emily O’Heir, Melanie C O’Leary, Ikeoluwa Osei-Owusu, Katrin Õunap, Lynn Pais, Sander Pajusalu, Alicia Pham, Eric A Pierce, Emma Pierce-Hoffman, Gianina Ravenscroft, Tony Roscioli, Vijay G Sankaran, Jillian Serrano, Elliott H Sherr, Shirlee Shril, Moriel Singer-Berk, Hana Snow, Volker Straub, Derek Tai, Tiong Y Tan, Ana Töpf, Ehsan Ullah, Grace VanNoy, Ivo Violich, Mark Walker, Susan M White, Monica H Wojcik, Emma Mitchell, Aziz M Al’Khafaji, Sheila Dodge, Kiran Garimella, Niall J Lennon, Stacey B Gabriel, Karen H Miga, Benedict Paten, Heidi L Rehm, Anne O’Donnell-Luria, Harrison Brand, Michael E Talkowski

## Abstract

Rare diseases collectively affect 1 in 10 individuals, yet current genetic testing fails to identify a causal variant for most cases. At present, cytogenetic methods and/or sequencing approaches such as exome (ES) or short-read genome sequencing (srGS) represent the state-of-the-art for comprehensive clinical discovery of sequence and structural variants (SVs), including copy number variants, balanced SVs, complex SVs, and tandem repeats (TRs). Recently, long-read genome sequencing (lrGS), coupled with multiomics data, has presented great promise to resolve variation in genomic regions recalcitrant to characterization by srGS such as highly repetitive simple repeat sequences and segmental duplications. However, there are few guidelines to enable clinical interpretation of genetic variation in these highly repetitive genomic regions, and the enthusiasm of the field in adopting lrGS has made it difficult to assess the true added diagnostic yield of this technology due to widely variable and inconsistently applied analytic pipelines and variable degrees of pre-screening by ES or srGS. Here, we investigated the contribution of SVs to rare diseases using srGS as a front-line strategy when paired with highly sensitive SV discovery and evaluate the added diagnostic yield of incorporating lrGS for a subset of cases. Our srGS analysis encompassed 1,462 families (3,450 individuals) recruited through the Broad Institute Center for Mendelian Genetics and the Genomics Research to Elucidate the Genetics of Rare Diseases (GREGoR) programs. Diagnostic SVs were identified in 5.4% of cases (79/1,462), of which 80% were uniquely detectable by srGS compared to standard cytogenetic techniques. For 96 families (including 10 families with a heterozygous variant observed in a known recessive gene of clinical relevance), we performed lrGS with methylation profiling, as well as long-read transcriptomic analyses in a subset of 20 trios. Analyses with lrGS yielded over 25,000 SVs per genome, 63% of which were not captured by srGS, along with an additional ∼200 rare SNV/indels per genome not previously captured and 12 differentially methylated regions per genome. Among these, we identified only one diagnostic variant not interpreted by srGS, an apparently mosaic *de novo* SNV in *CASK* that was absent in the srGS callset due to allelic imbalance. No new diagnoses were supported by long-read transcriptomics or episignatures. In this well characterized rare disease cohort, the added diagnostic yield was thus 1.04% (1/96 families). Following a systematic literature review of prior lrGS studies, we find that most reported diagnoses were detectable by srGS and that our added diagnostic yield is consistent with those prior studies. These studies emphasize the significant impact of comprehensive SV discovery in rare disease cases and further demonstrate the power for increased discovery of novel genomic variation and episignatures from lrGS. Nonetheless, they also serve to temper expectations of dramatic diagnostic advances in rare disease patients until there is more extensive annotation of the functional and clinical impact of all coding and noncoding variation uniquely accessible to lrGS with extensive reference databases spanning highly repetitive genomic sequencing that could be enabled by this transformative technology.

## INTRODUCTION

Rare diseases collectively impact millions of individuals worldwide, presenting a substantial burden on affected families and healthcare systems.^1^ To date, more than 8,000 rare diseases have been reported, with estimates suggesting that up to 80% of cases have a genetic basis.^2^ Rapid advances in genomic sequencing technologies over the past decade have enabled the adoption of exome sequencing (ES) or short-read genome sequencing (srGS) as a potential front-line strategy for rare disease diagnosis, yet a majority of patients still lack a known genetic etiology following evaluation with these technologies.^3,4^ Multiple factors underlie the challenges associated with undiagnosed rare disease cases, including polygenic contributions of common variants to developmental disorders,^5–10^ incomplete functional interpretation of noncoding regulatory variation in the human genome,^11^ and the technical challenges of reliable identification of structural variants (SVs) localized to highly repetitive genomic regions.^12^

The role of SVs has traditionally been under-surveyed in studies of rare diseases and trait associations. Research studies initially focused on the discovery and interpretation of far more abundant classes of single-nucleotide variants (SNVs) and small insertions/deletions (indels), supplemented with large copy number variants (CNVs) identified by microarray and/or targeted panel sequencing. Many prior ES and srGS studies have either primarily studied SNVs/indels or restricted SV detection to a small set of algorithms.^13–15^ While new algorithms and analytic frameworks have facilitated the rapid and systematic discovery of CNVs in ES data,^16,17^ as well as all classes of SVs (deletions, duplications, insertions, translocations, inversions, and complex SVs) from srGS across large cohorts,^18^ these approaches remain inconsistently applied, thus leaving systematic holes in understanding the complete contribution of SVs to rare disease.^3,4,13,16,19^

Recently, long read genome sequencing (lrGS) has emerged as a technology with the potential to close many gaps in the genetic diagnosis of rare disease. It enables near complete ascertainment of genomic variation through alignment-based analyses and *de novo* assembly. The primary discovery power of lrGS lies in the ability to capture variants that reside in highly repetitive regions that have traditionally been recalcitrant to srGS discovery such as tandem repeats (TRs), pseudogenes, segmental duplications, and low-complexity regions.^20–25^ The capacity to simultaneously survey multiomics information is also a major advance as lrGS technologies provide access to DNA methylation profiling and haplotype-specific methylation signatures.^26–28^ The greatest value in lrGS is evidenced in SV discovery, as most lrGS analytic methods to date detect approximately twice as many SVs as srGS and resolve a broader spectrum of repeat-mediated variants at higher precision.^22–25^ Despite this promise, the true added diagnostic yield of lrGS relative to existing technologies remains unclear. Both srGS and lrGS technologies show high concordance in coding, unique, and repeat-masked sequences in the human genome when comprehensively analyzed and compared, and these genomic regions harbor most known pathogenic variants from OMIM, suggesting the gain from lrGS in genetic diagnostics could be marginal and is fully dependent on variant classification evidence in the field.^24^ Indeed, significant inconsistencies in study design have produced divergent interpretations of diagnostic yields. Due to the high costs of lrGS, many studies have preselected individuals based on phenotype or with unresolved variants detected with lower resolution technologies; moreover, the analytic challenges of lrGS have restricted some studies to analyses of targeted loci rather than exploration of the complete spectrum of accessible variation, while still other studies have not performed benchmarking against current state-of-the-art srGS analyses and instead compared lrGS to ES or targeted sequencing.^29–34^ Incomplete SV discovery in srGS is also a pervasive challenge in rare disease diagnostics and leads to inflated estimates of the added yield of lrGS as technology-specific rather than due to analytic gaps in the first-line analysis.^35–37^ Of course, genetic architecture of the phenotypes selected is also a major driver of diagnostic yield, as individuals with severe and syndromic developmental phenotypes are more likely to harbor a dominant-acting high impact variant than those with more subtle or complex traits such as autism.^6,19,38^ A rigorous and unbiased comparison is therefore essential to define the current utility of lrGS.

In this study, we sought to comprehensively assess the impact of SVs on rare diseases through a systematically collected long-term study of Mendelian disorders using srGS as a front-line strategy in 3,450 individuals from 1,462 families with suspected monogenic diseases, recruited at the Broad Institute as part of its Center for Mendelian Genomics (CMG), a research center within the National Human Genome Research Institute’s (NHGRI) Genomics Research to Elucidate the Genetics of Rare Diseases (GREGoR) Consortia.^11,39^ We next evaluated the ability of lrGS to identify novel diagnostic variants, epigenetic signatures, and differentially methylated regions (DMRs).^40–45^ In a subset of 20 families, we also explored the added value of long-read RNA sequencing (lrRNAseq) for full-length isoform sequencing and quantification to clarify the impact of splice-altering variants of uncertain significance in undiagnosed rare diseases.^46–49^ Our results confirm the profound impact of SVs underlying rare disease diagnoses and demonstrate the intriguing discovery power of lrGS, but the negligible added diagnostic value in this cohort illuminates the critical new for new metrics and functional annotations to facilitate interpretation of genomic sequences not previously accessible in the field of clinical genomics.

## RESULTS

### Cohort and technology overview

#### Short-read genome sequencing (srGS)

We performed srGS on 3,450 individuals from 1,462 rare disease families, including 111 quartet families (proband, affected sibling, and both parents), 644 trios, 154 duos, 518 singletons, and 35 families with other structures (Figure 1A). Sequenced individuals were largely balanced for sex (1,740 females and 1,710 males). The primary indication for genetic testing spanned neurodevelopmental (27%), neuromuscular (25%), ophthalmological (10%), immune (7%), metabolic (5%), hematological (5%), genitourinary (3%), cardiovascular (2%), musculoskeletal (1%), and other syndromic as well as non-syndromic disorders (15%) (Figure 1B). Phenotypes were recorded as HPO terms (range 1-43 terms per proband, median=3 terms). At least 748 families have undergone prior genetic analysis reported in previous studies.^3,16,50–61^

**Figure 1.**
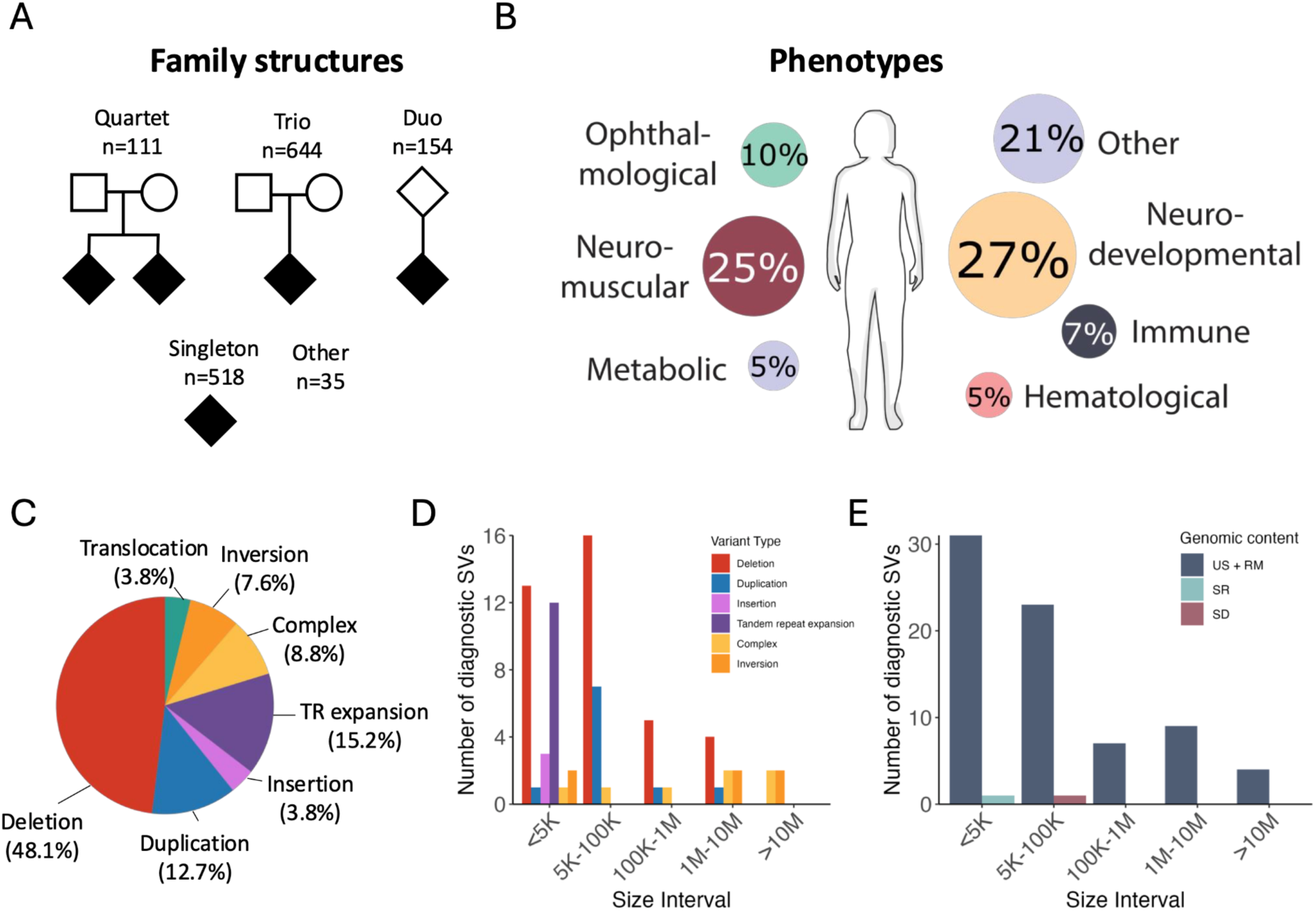
Cohort description, phenotypes and diagnostic SVs identified by srGS. A) Family structure counts. B) Phenotype proportions across the cohort. C) Overall proportion of diagnostic SVs by variant type. D) Diagnostic SVs by variant type and size interval; translocations excluded. E) Diagnostic SVs by genomic content and size interval; translocations excluded. TR=tandem repeat; RM=repeat masked sequence; SD=segmental duplications; SR=simple repeats; US=unique sequence. US+RM are regions of the genome that are largely accessible and accurately aligned with srGS. SD+SR are highly repetitive sequences that are more accessible to lrGS than srGS.

#### Long-read genome sequencing (lrGS)

We performed both srGS and lrGS on 242 samples from 96 families including use of the Pacific Biosciences platform on 181 individuals and ONT platform on 61 individuals. During the course of analyses, 14 of these families were ultimately solved with srGS, while 82 remained unsolved. Notably, among the 96 families, 10 were chosen due to the presence of a heterozygous SNV/indel in a recessive disease gene to explore the likelihood of a cryptic SV or other variant uniquely detectable by lrGS on the other allele.

#### Short-read RNAseq (srRNAseq)

srRNAseq was performed on 273 cases, 9 cases of which were previously known to harbor a diagnostic SV (see Methods); this technology was used to confirm functional disruption of predicted genes in these cases.

#### Long-read RNAseq (lrRNAseq)

We performed PacBio Kinnex sequencing for 60 individuals from 20 trio families sequenced with lrGS, with samples selected based on availability and quality of RNA (see Methods).

### Discovery of pathogenic SVs and TRs by srGS

We evaluated SVs and TR expansions in 1,462 rare disease families. SV-based diagnoses were detected in 5.4% of families (79/1,462) and included 3.4% (49/1,462) dominant/biallelic recessive acting SVs, 1.2% (18/1,462) SVs that were compound-heterozygous in trans with an SNV/indel in a recessive disease gene, and 0.8% (12/1,462) TR expansions (Table S2). The majority of diagnostic SVs were deletions and duplications, accounting together for 60.8% of cases (48/79), followed by TR expansions (15.2%, 12/79), complex SVs (8.8%, 7/79), inversions (7.6%, 6/79), translocations (3.8%, 3/79), and insertions (3.8%, 3/79) (Figure 1C). Excluding translocations, most SVs were smaller than 100 kbp (73.7%, 56/76), falling below the resolution threshold of standard cytogenetic diagnostic technologies such as chromosomal microarray or karyotyping (Figure 1D). The SV diagnostic rate was highest among families with metabolic (10%, 8/80), and neuromuscular (7.9%, 14/178) phenotypes (Figure S1A). Families where data from both biological parents were available for variant phasing had a higher diagnostic rate (6.7%, 51/766) versus those where only one or neither parent was available (4%, 28/696) (Fisher’s exact test p=0.0278) (Figure S1B). In addition, 13 SVs were classified as strong candidates but did not rise to the classification of pathogenic or likely pathogenic (LP), increasing the possible SV yield to 6.3% (Table S3).

Notably, 98% (76/79) of diagnostic SVs were identified in genomic regions readily accessible to srGS (unique or repeat-masked sequences), as these regions harbor most protein coding sequence in the human genome, while only three clinically interpretable SVs were localized to highly repetitive segmental duplications (SD) or simple repeat (SR) sequences (Figure 1E). These findings have implications for the value added of lrGS, were the vast majority of novel variation discovered by this technology and missed by srGS span these highly repetitive SD/SR regions that represent approximately 11% of the human genome.^24^ In addition, genomic disorder (GD) SVs, which are disease-associated recurrent CNVs caused by non-allelic homologous recombination (NAHR), were identified in five cases: three were diagnostic (a 16p12.2 deletion, a 5p15 deletion, and a 15q11-q13-BP1-BP3 deletion), and one was probably diagnostic (a 2q11.2 deletion) (Table S2). In one family, we identified a 16p11.2 BP4-BP5 duplication (RGP_1526) that partially explained the phenotype of the proband (microcephaly and seizures), inherited from the unaffected father.

Among the 79 clinically relevant SVs, seven complex SVs were identified. i) A duplication-inversion-duplication affecting *ANKRD26* was identified in family ABC012, a large pedigree comprising nine individuals across three generations affected by maternally-inherited thrombocytopenia.^50^ ii) A *de novo* complex SV was observed in family HK029 consisting of a 13 kb deletion and a 7 kb tandem duplication impacting *MED13L*, with a phenotype of mild intellectual disability, motor developmental disorder, and blepharophimosis. iii) A hemizygous duplication-inversion-deletion affecting *DMD* was discovered in an individual in family HQ diagnosed with Duchenne muscular dystrophy. iv) In two affected siblings (family EA) with muscular dystrophy, a hemizygous complex rearrangement was resolved that consisted of a 116 kb duplication from chromosome 8 inserted into intron 43 of the *DMD* gene on the X chromosome. Additional complexity included a duplicated 124 bp segment of intron 43 (chrX:32276895–32277018) flanking the chromosome 8 insertion, along with a 13 bp insertion (GCCTTTGCCCACA) adjacent to the duplicated segment. v) A *de novo* complex SV comprising two duplications (576 kb and 112 kb) and one deletion (93 kb) was reconstructed in the proband (RGP_1786). This rearrangement resulted in a full copy gain of *CSRNP3* and *SCN2A*, a partial duplication and deletion of *CHN1*, and duplication of the transcription start site of *SCN3A*. The proband presented with benign early-onset infantile seizures, a phenotype previously associated with *SCN2A* duplications,^62^ and the variant was therefore classified as partially explanatory of the phenotype. vi) A homozygous duplication of exon 13 of *PLEKHG5* was identified to be inverted and inserted near exon 16, in an individual with a neuromuscular disorder (RGP_921). vii) A highly complex *de novo* SV was found in the affected member of family RGP_563 presenting with syndromic intellectual disability that involved 37 breakpoints, 10 deletions, and 79 genes, collectively accounting for a rearrangement that spanned 16 Mb, all fully resolved by srGS (Figure 2A, Table S4).

**Figure 2.**
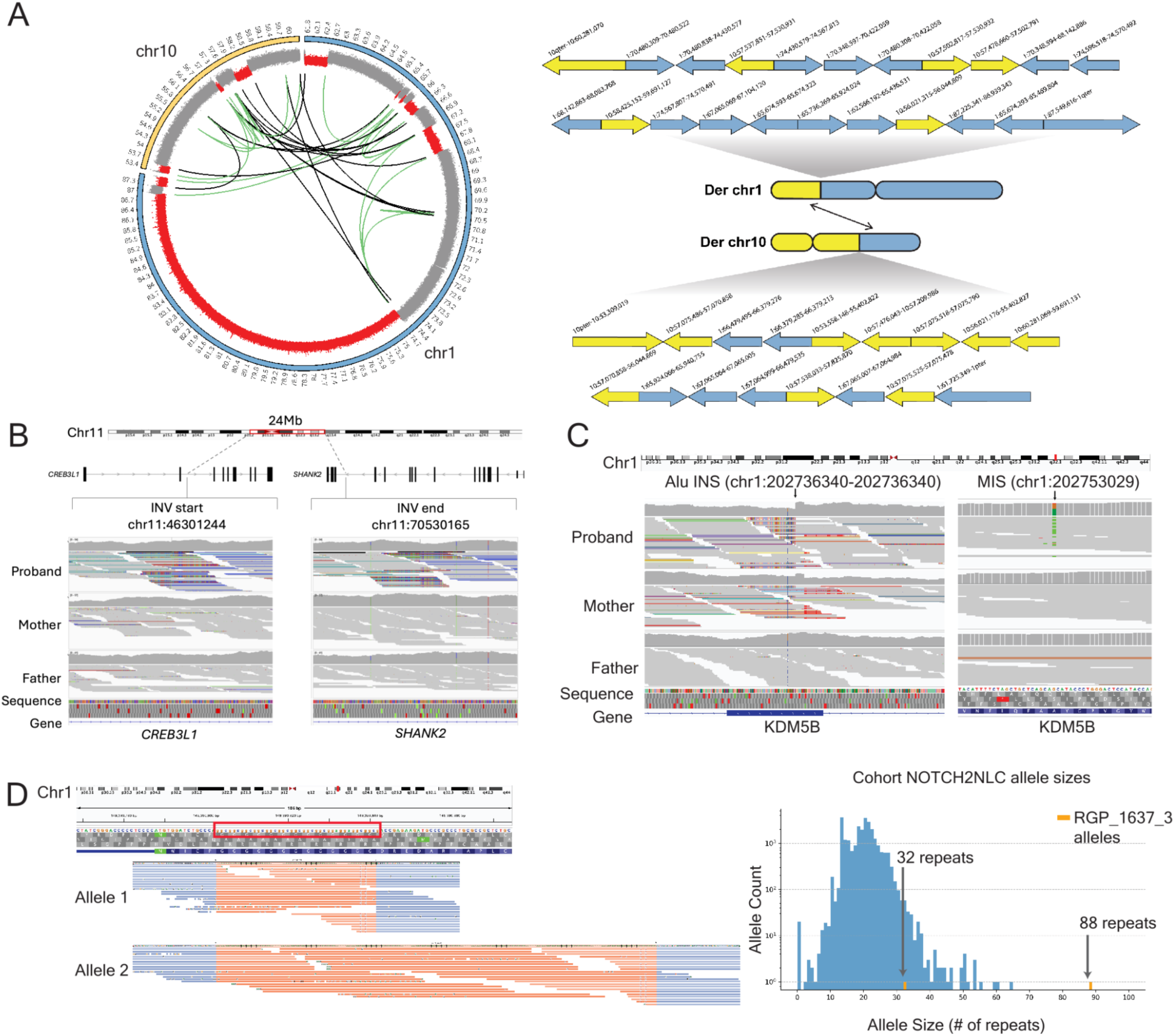
Examples of pathogenic SVs identified in this cohort. A) Highly complex *de novo* SV involving 37 breakpoints with shattered segments between chromosomes 1 and 10 in a patient with dysmorphic features and a neurodevelopmental disorder. Der=derivative. B) *De novo* inversion predicted to disrupt SHANK2 in a case with intellectual disability. INV=inversion. C) Alu insertion in KDM5B identified in trans with a *de novo* missense variant in a case with Lennox-Gastaut syndrome. INS=insertion; MIS=missense D) TR expansion in the RGP_1637 family involving NOTCH2NLC gene. #=number.

There were multiple mechanisms involved in diagnostic SVs that contributed to affected individuals’ phenotypes. Five of the 79 diagnostic SVs were larger than 10 Mb, including three inversions, one deletion, and the complex SV in RGP_563 (described above). The largest SV was a 24.23 Mb *de novo* pericentric inversion, which was a balanced event predicted to disrupt *SHANK2* in an individual (ROS_28) with a neurodevelopmental disorder (Figure 2B). There were 18 families with diagnostic SVs that represented compound heterozygous events in combination with a SNV/indel, and we were able to phase the variants’ haplotypes from srGS in 13 of the cases. One example is family RGP_1646, diagnosed with Lennox-Gastaut Syndrome. In this case, ES alone discovered a *de novo* missense variant in *KDM5B*, while subsequent srGS due to suspicion of recessive disease also detected this variant as well as a maternally inherited Alu insertion in trans (Figure 2C). In 12 additional families, the causal variant was a TR expansion involving a total of eight genes (Table S2; *ATXN1, ATXN2, CNBP* [x2], *DMPK* [x3], *EP400, NOP56, NOTCH2NLC, RFC1* [x2]). One such example was at the *NOTCH2NLC* locus in an individual with progressive muscle weakness (RGP_1637) (Figure 2D). Expansions at this locus to 66 or more GGC repeats have been associated with neuronal intranuclear inclusion disease (NIID)^63^. In this case, a 32/88 repeat expansion was initially identified in the srGS data and was confirmed to be a 32/145 repeat by fluorescence amplicon length analysis (Figure S2).

### Transcriptomics support annotation of the functional impact of SVs

Nine of the probands harboring clinically relevant SVs had available srRNAseq, which we used to confirm functional disruption of predicted genes in seven of the nine probands (Table S5). In one family, BEG_1025, srRNAseq supported reclassification of a noncoding SV from a variant of uncertain significance (VUS) to pathogenic (P). In this family, the affected male harbored a noncoding SV representing a hemizygous intronic L1HS insertion in intron 3 of *MTM1* on chrX with uncertain functional impact (Figure 3A-C). srRNAseq identified a complex splicing pattern characterized by reduction of transcripts with the exon 4-5 junction and novel junctions between exons 3-5, 3-6, and 1-7 (Figure 3A-C), all predicted to result in nonsense mediated decay, with gene expression outlier analysis (OUTRIDER) demonstrating significant underexpression outlier of *MTM1* (Z < -7; p < 10^-9^) compared to a cohort of muscle RNA-seq from 384 individuals with rare disease (Figure 3A-C), supporting a genetic diagnosis of *MTM1*-related myotubular myopathy. Notably, the SV was detected in the unaffected mother in only 7% (4/54) of reads, indicating a likely mosaic event that has implications for genetic counseling around recurrence risk. Among the two probands in whom the causal SV did not result in an expression outlier, one (family RGP_94) harbored a *de novo* deletion of an lncRNA (*CHASERR*) that displayed a regulatory effect resulting in transcript allelic imbalance and increased protein abundance of CHD2, a disease mechanism previously reported.^56^ The second case (family EA) harbored a hemizygous, likely mosaic complex SV involving a 116 kb duplication of chr8 inserted into intron 43 of *DMD* on chr X, described in more detail above. srRNAseq confirmed that the chromosomal fusion generates multiple novel splice junctions between chrX and chr8, while retention of wild-type splicing in an expected number of reads further supports somatic mosaicism (Figure S3).

**Figure 3.**
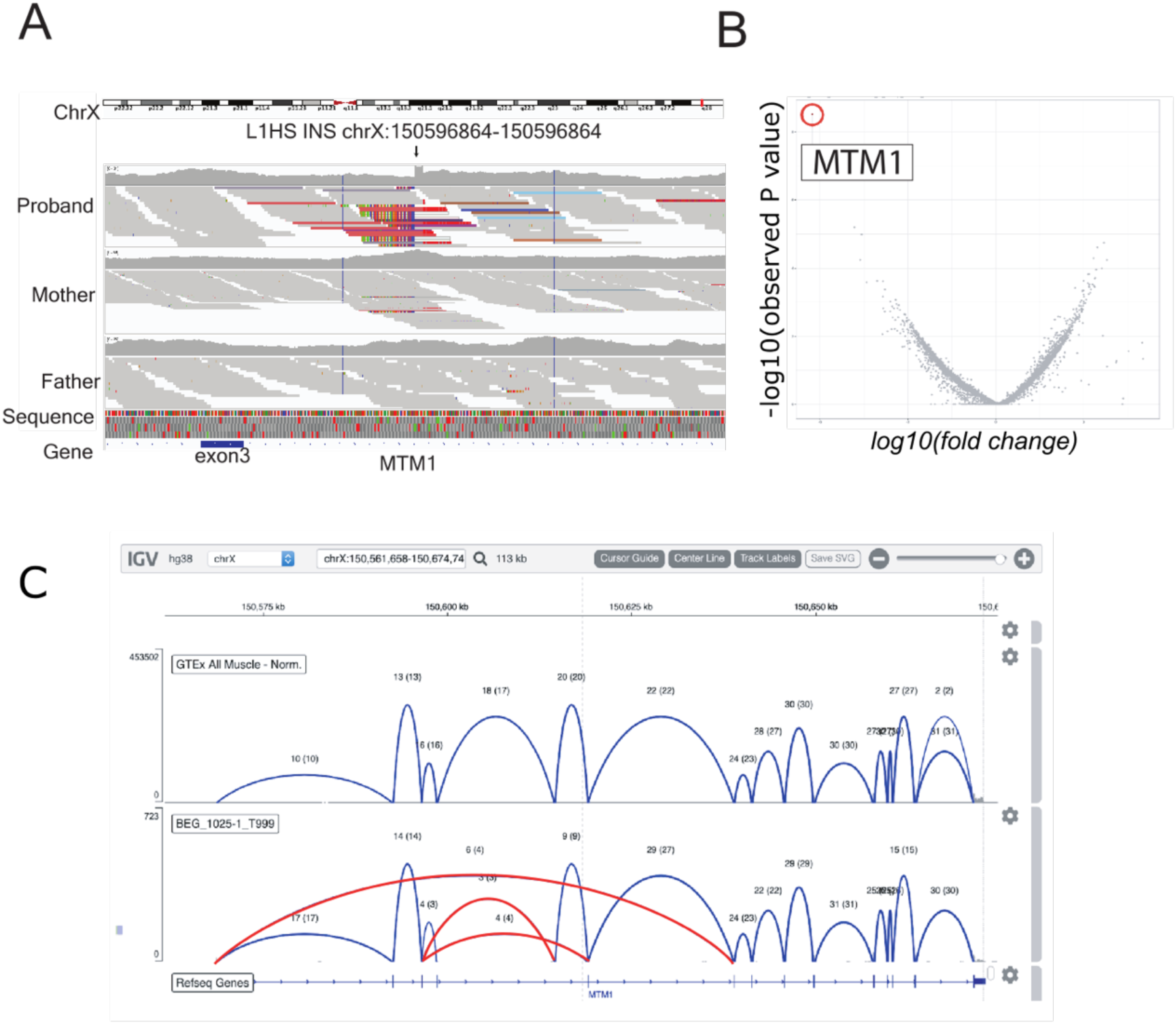
RNA-seq supports disruption of *MTM1*. A) Alignment IGV plot of the LINE1 (L1HS) insertion identified in BEG_1025 family shows the variant is hemizygous in the proband and likely mosaic in the mother, with four reads supporting the insertion. INS=insertion. B) Volcano plot of gene expression analysis by OUTRIDER demonstrating a significant underexpression outlier for only one gene, *MTM1* (Z-score < -7; p < 10^-9^) compared to a rare disease cohort of 384 muscle RNA-seq samples. C) Sashimi plot demonstrating aberrant splicing patterns (novel junctions in red, known junctions in blue) in patient (bottom) in comparison to GTEx muscle controls (top).

### Limited added diagnostic value from long-read sequencing in this rare disease cohort

To explore the added diagnostic value of lrGS, 242 samples from 96 families were sequenced with both srGS and lrGS using ONT (n=61 individuals from 20 families) or Pacific Biosciences (n=181 individuals from 76 families). The lrGS data produced an average genome-wide coverage of 30x (range 11-55x) and an N50 of 20.2 kb (range 11-45 kb), with a median read identity aligned to the GRCh38 reference genome of 99.7%. Compared to srGS, improved coverage was seen in lrGS for 479 challenging clinically significant genes (Supplemental Data; Figure S7A-C).^64^

For SNV/indels, lrGS detected a median of 203 high-quality (GQ ≥ 30, allele balance [AB]≥0.2) rare variants per proband that had not been detected at high quality in the corresponding srGS sample, of which a median of 23 per sample were predicted damaging (VEP^65^ moderate/high impact) and a median of four per sample impacted disease-associated genes, which were manually reviewed for phenotype, inheritance, and disease mechanism consistency to determine diagnostic relevance. For SVs, we identified a total of 28,697 distinct rare (AF <= 0.1 compared to both srGS and lrGS-based reference catalogs; see Methods) variants from lrGS, with a median of 757 per sample. Comparing to the matched srGS data, 87% of srGS SVs per genome were detected in lrGS, suggesting high precision of the srGS SV calls. By contrast, and consistent with previous studies,^24^ only 28% of the lrGS calls were discovered in srGS and the vast majority (86%) of these lrGS-unique SVs localized to tandem repeat sequences (Table S6). A total of 94 lrGS SVs (median 4 per sample) were predicted as LoF, intragenic exon duplication, or full copy-gain of a protein-coding gene and were reviewed for diagnostic relevance as described above. All rare lrGS SVs were reviewed for diagnostic relevance in combination with small variants to identify candidate compound heterozygous diagnoses (see Methods and Table S1).

Among the 96 probands, 15 harbored a diagnostic variant (Table S7); 14 were observed by both srGS and lrGS, while one causal variant (a predicted mosaic SNV) was uniquely discovered by lrGS (1/96 probands, added diagnostic yield of 1.04%; Table S1, Figure 4A-B). Confirming the power of re-analysis, the largest fraction of new diagnoses compared to the first-pass srGS analysis were derived from novel gene reports published over the course of the study. Six of the 15 variants involved the report of new gene-disease relationships, including the noncoding gene *RNU4-2*, which is now recognized as a frequent cause of the neurodevelopmental ReNU syndrome (MIM: 620851).^66^ One variant detected by both technologies required functional evidence from a high throughput splicing assay (HTSA)^67^ to upgrade a noncoding VUS to LP. In family RGP_1786, a complex SV was resolved by both technologies (formed by a partial duplication of *SCN3A*, inversion duplication of *SCN2A* and *CSRNP3* and deletion of *CHN1*). Surprisingly, among the 10 families that were selected due to a single candidate variant in a recessive disease-associated gene to explore potential novel variants observed in trans, no compound heterozygous variants were identified in any families.

**Figure 4.**
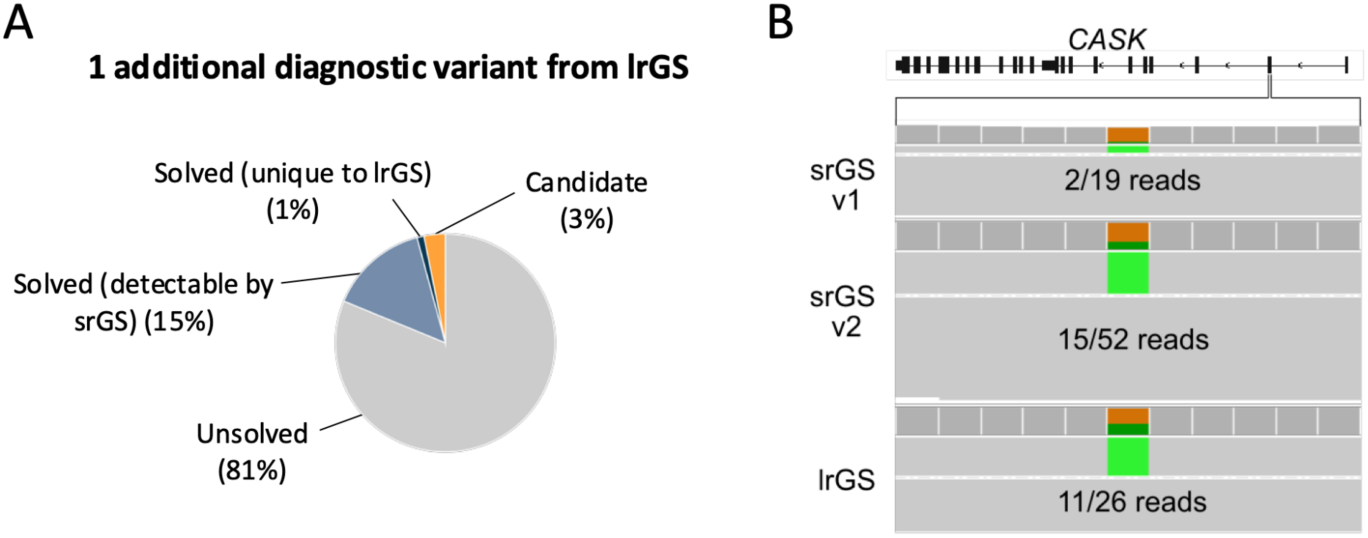
Added diagnostic yield from lrGS. A) Number of candidate variants identified in the lrGS families by family solve status. C) IGV snapshot of the nonsense SNV in *CASK* gene, the only variant exclusively identified by lrGS (in the RGP_1777 family). Alternate supportive reads are shown in green. The variant was absent in the original srGS (srGS v1) due to stochastic loss of the alternative allele, but present in a repeated srGS (srGS v2) and the lrGS.

Only one variant uniquely observed by lrGS was determined to be diagnostic, resulting in a 1.04% added diagnostic yield (Table 1, Figure 4A-C). This variant was a ClinVar 2-star pathogenic nonsense variant in *CASK*, consistent with the proband’s (RGP_1777_3) phenotype of intellectual disability, global developmental delay, and microcephaly. It was present in 11 of 26 reads (allele balance 42%) in the lrGS, while post-hoc manual review found the variant was present at low allele balance but filtered in the srGS data (2 of 19 reads, 11%; Figure 4C). Coverage of this chrX position by srGS does not suggest a systematic coverage deficiency, with mean coverage of 38x in females (among 85 females with both srGS and PacBio lrGS), despite being only 19x for this female proband. The variant is not in a GC-rich or low complexity region, and *CASK* is not listed among the potential clinically relevant genes that might be challenging to sequence by srGS.^64^ Resequencing of a new srGS library with the same DNA sample as used in the PacBio sequencing yielded the SNV in 15/52 reads (allele balance of 29%; Figure 4C) further supporting a case of mosaicism. We further validated this result by PCR amplification and Sanger sequencing of the DNA sample from blood and buccal swab, both demonstrating reduced signal of the alternate allele compared to expectation of a true heterozygous state (Figure S4A-B). Consistently, the patient presented with a relatively mild phenotype compared to that typically described for patients with a pathogenic *CASK* variant, further consistent with mosaicism.

Aside from the minimal number of new diagnoses, lrGS demonstrated a number of other advantages over srGS. In several cases with diagnostic SVs, lrGS provided improved breakpoint resolution, largely driven by its ability to better resolve SVs mediated by homologous mobile elements at the breakpoints (four by Alu elements and one by LINE-1 elements) (Figure S5A-F). For example, one LINE-1-mediated SV, a 1.1 Mb deletion spanning multiple genes including *CNOT2*, was detected in RGP_2210_3 by srGS using depth-based methods, while the lrGS data assembled the breakpoints to a 200 bp region inside the LINE-1 element (Figure S5A). In another case, lrGS phased two variants in *HECTD4* spanning greater than 65 kb of sequence using only the proband’s reads to confirm that a *de novo* coding deletion and an inherited SNV at a splice donor site occurred in trans (Figure S6A). The family was sequenced as a trio by srGS, so phasing and interpretation were already possible, but lrGS demonstrated the potential for single-sample assembly and phasing to unambiguously derive compound heterozygous events. In addition to the diagnostic SVs described above, lrGS analysis detected a candidate highly complex mosaic rearrangement in individual RGP_1316 (Table S8). The variant was predicted from srGS with the detection of five large *de novo* duplications across four different chromosomes, but the larger structure and substantial additional complexity was resolved by manual review of the ONT data. The initial SV detection algorithms found no additional breakpoint complexity, but using bigclipper^26^, a tool we previously developed to find candidate rearrangement breakpoints by identifying pile-ups of clipped reads (see Methods), we resolved additional *de novo* breakpoints that were unique in the lrGS data.^26^ Overall, this analysis resolved to a mosaic chromoanasynthesis event involving at least ten chromosomes, 24 distinct breakpoints, and nine duplicated regions spanning ∼8.8 Mb, some of which spanned complex nested duplications (Figure S6B). The results are highly suspicious of a clinically relevant rearrangement. srRNAseq detected three gene expression outliers and outlier splice junctions in 46 genes impacted by the cxSV, including *CARS2*. However, despite the resolution enabled by lrGS and the clear functional impact from srRNAseq, this event could not be considered diagnostic as it was mosaic in blood, with roughly ∼20% of reads supporting it, and the affected genes were not clearly linked to the proband’s phenotype.

Finally, we searched for potential diagnostic variants in complex and repetitive loci made accessible by lrGS. To query clinically relevant genes with highly similar paralogs, we applied the tool Paraphase and analyzed results for 14 clinically-relevant regions of the genome (see Methods).^68^ In RGP_1556, a proband with congenital adrenal hyperplasia and a maternally inherited pathogenic SNV in *CYP21A2*, Paraphase confirmed the presence of a p.Val282Leu pathogenic variant on the maternal copy of the RCCX locus (known from srGS), as well as a benign pseudogene duplication in cis. The paternal allele was found to only have a benign pseudogene deletion, with no evidence of a second pathogenic event to complete the diagnosis. Moreover, RNA-seq indicated normal expression of both alleles. This case therefore remains unresolved. After TR genotyping of 71 known disease-associated loci using TRGT,^69^ no diagnostic or high-interest candidate TR expansions were identified in either the PacBio or ONT datasets.

Discovery of differentially methylated genomic regions and disease episignature concordance We pursued lrGS-derived DNA methylation analyses across the 242 samples, including analysis of known imprinting disorders, detection of outlier differentially methylated regions, and matching of episignatures to known disorders. We ran NanoImprint^70^ but did not detect any evidence of alterations associated with known imprinting disorders. Using the BSmooth package^71^, we detected a median of 12 outlier differentially methylated regions (DMRs) per sample at CpG islands or cCREs (candidate cis-regulatory regions)^72^, and a median of 4 such DMRs per sample overlapping promoters, enhancers, or the gene body of disease-associated genes (Figure 5A), though no DMR-associated genes were significant expression outliers in whole blood srRNAseq. A total of 57 DMRs overlapped with a rare SNV/indel/SV; however, clinical review did not identify any strong phenotype matches among these. We further analyzed 36 episignatures^44,73^ derived from established methylation disorders using the NSBEpi method.^74^ We found three episignatures to yield a high number of apparent false positives in unaffected parents or probands with unrelated phenotypes (Tatton-Brown-Rahman syndrome, Cornelia de Lange syndrome, and *CHD8* episignatures), while two weakly positive results for additional episignatures were observed in unaffected parents.

**Figure 5.**
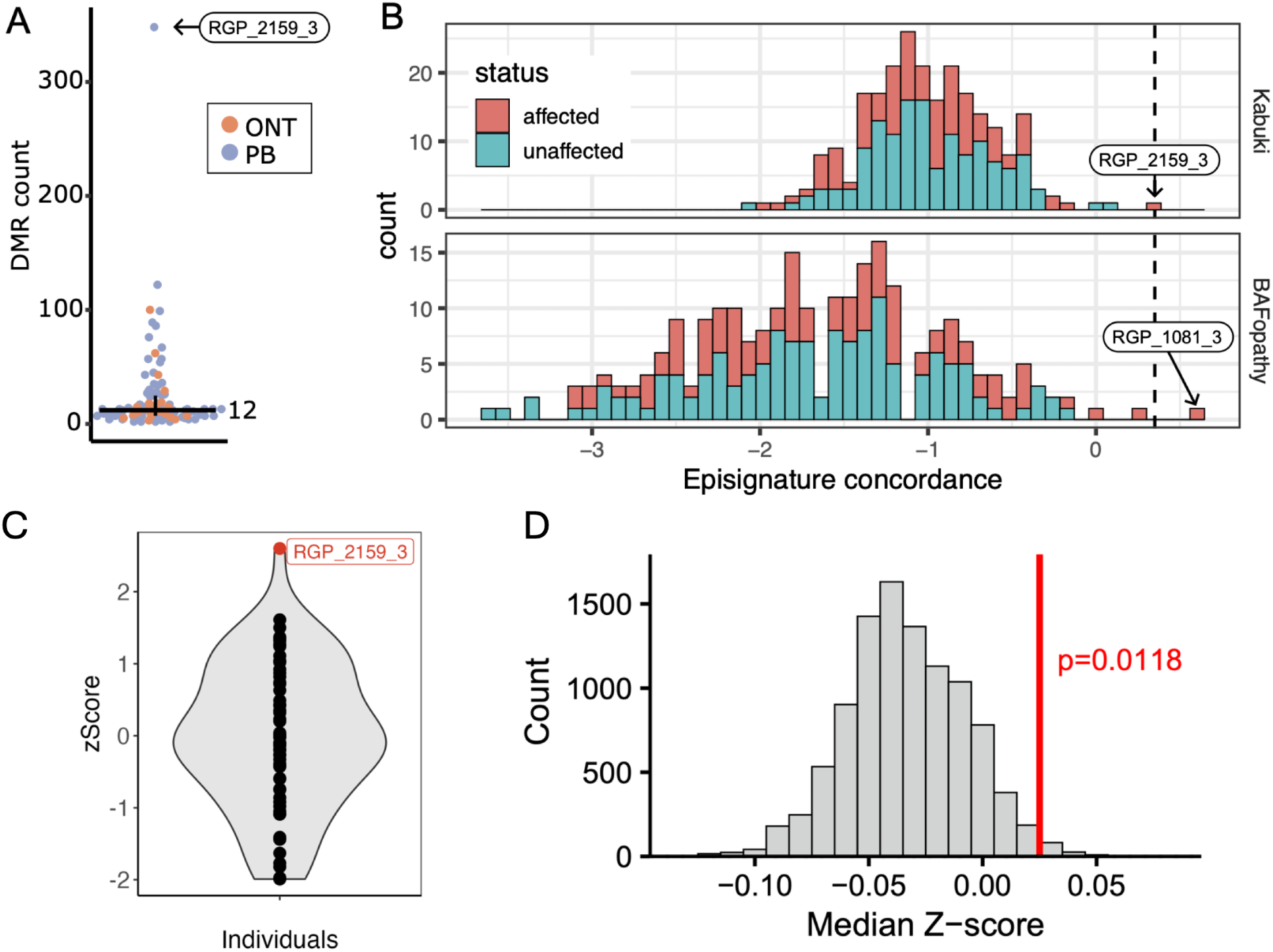
Methylation and expression analyses from long-read sequencing. A) Differentially methylated regions (DMR) counts per sample, colored by technology. The horizontal bar represents the median value across samples. Extreme outlier sample is labeled. B) Episignature concordance for Kabuki syndrome (top) and BAFopathy (bottom) across the cohort, with affected or unaffected status indicated. The dotted line represents the threshold (as recommended by Geysens et al.^74^) for a match. C) OUTRIDER-calculated Z-scores of MAZ gene for the lrRNAseq samples. D) Median Z-score permutation analysis for DMR of proband.

Among affected probands, two samples had episignature scores of interest (Table S9). Proband RGP_2159_3 was clinically suspected of having a disorder disrupting methylation, either CHARGE syndrome (causative gene *CHD7*) or Kabuki syndrome (causative genes *KMT2D/KDM6A*) based on clinical phenotype. This case had a high number of DMRs (n=324, while the cohort median was 12; Figure 5A), and a mild Kabuki episignature concordance (score of 0.32, just above the 0.3 cutoff; Figure 5B), consistent with a mildly abnormal Kabuki methylation signature reported by clinical testing. The case harbored a *de novo* SNV in the *MAZ* gene that was classified as a VUS as the gene is not currently disease-associated. The *MAZ* protein has previously been shown to colocalize with CTCF, interact with the cohesin complex component RAD21 (in which pathogenic variants can cause Cornelia De Lange Syndrome) and contribute to gene expression and genomic architecture during development.^75,76^ *MAZ* is also part of the well-established 16p11.2 reciprocal genomic disorder locus.^8,77–80^ Given that episignatures have been defined for 16p11.2 deletions and *CHD8* variants,^73^ we evaluated these episignatures across all cases in our cohort, and RGP_2159_3 did not show concordance with either episignature (Figure S8). This variant thus remained classified as VUS. The second case of interest involved proband RGP_1081_3, which scored 0.6 (well above the 0.3 cutoff) for concordance with the episignature of Coffin-Siris and Nicolaides-Baraitser syndromes, which are caused by pathogenic variants in subunits of the BAF chromatin remodeling complex, and commonly referred to as BAFopathies (Figure 5B). RGP_1081_3 has a broadly consistent phenotype with a BAFopathy, including autism (non-verbal), global developmental delay, focal-onset seizure, infantile spasms, congenital umbilical hernia, gastroesophageal reflux disease and myopic astigmatism. However, we were unable to identify potentially causal variants among BAF complex genes and 200 genes implicated in human DNA methylation, nor expression or splicing outliers among a subset of these genes that are well expressed in blood. In an additional sample of interest, proband RGP_1012 with an overgrowth syndrome, we detected a rare (absent in gnomAD) noncoding *de novo* variant overlapping a predicted enhancer in NSD1 (chr5:177132105C>A), a gene associated with Sotos syndrome and overgrowth, consistent with the proband’s phenotype; however, both our episignature analysis and confirmatory clinical testing were negative for Sotos and NSD1 concordance. Overall, the methylation and episignature data yielded intriguing new insights and analyses represented a tractable number of novel findings to review, but no additional diagnoses were derived from these analyses of the lrGS methylation data.

### Long-read RNA sequencing performed in 20 trio families

We further sought to identify any transcriptomic aberrations using lrRNAseq from blood to prioritize candidate variants.^81^ For these analyses, 20 trio families with lrGS were selected for PacBio Kinnex sequencing. After identifying specific genes and variants of interest based on phenotype match and computationally predicted effect, lrRNAseq data were manually reviewed by identifying novel transcripts and junctions associated with the proposed gene or variant and performing manual inspection with IGV; no candidate genes or variants were associated with abnormal transcripts (Table S10). We next explored genes containing highly expressed novel transcripts that also contained either a *de novo* variant or at least one rare predicted LoF variant, with candidates selected for manual examination in IGV based on phenotype match; however, no plausible causal genes were identified. Given that proband RGP_2159_3 had lrRNAseq data available, we inspected the OUTRIDER Z-scores for the *MAZ* gene across all probands and identified that it was a positive expression outlier in this sample (Z=2.6) (Figure 5C). This result was significant for this gene (p=0.0034), but not in a genome-wide analysis after Bonferroni multiple testing correction (p=1). Furthermore, while srRNAseq analysis indicated that no DMR-associated genes were significant expression outliers, we found that DMR genes as a whole were more likely to be abnormally expressed: the median |Z-score| for the DMR genes in each proband was significantly higher than that of gene sets of comparable size (permutation p=0.0118) (Figure 5D), emphasizing the potential of multiomics to support the predicted functional effects of VUS.

### Diagnostic yield comparison to previously reported studies

Given the modest increase in diagnostic yield for variants requiring lrGS in our study (1.04% across 96 families), we compared these findings to previously published studies that included individuals who underwent both srGS and lrGS and focused on the subset of studies that clearly reported which variants were identified by each technology. Cohen et al.^37^ conducted both srGS and lrGS on 203 probands (analysis conducted on Table S6 of Cohen et al., estimated to represent 166 families after excluding related individuals as cross-referenced in Table S1 of Cohen et al.) and identified two families with P/LP variants uniquely detected by lrGS. These included a compound heterozygous deletion in trans with a SNV in *AARS2* (cases 110 and 111), and a TR expansion in *STARD7* identified in a large family pedigree (cases 188–193). These findings correspond to a 1.2% increase in diagnostic yield attributable to lrGS from that study. Hiatt et al.^36^ performed lrGS on 96 families with negative prior srGS results and identified two cases with P/LP variants exclusive to lrGS, representing a 2.1% (2/96) increase in diagnostic yield (Table 1 from Hiatt et al.): one involving a TR expansion in *PHOX2B* not detected by srGS, and another involving a mosaic SNV in *SHANK3* missed due to allele imbalance (only two supporting reads in srGS), comparable to our findings in *CASK* described above. All other P/LP variants identified by lrGS were accessible to srGS discovery, consistent with findings from our study. A recent study by Ek et al.^82^ performed srGS and lrGS in 100 unrelated individuals with neurodevelopmental or neuromuscular phenotypes. A genetic diagnosis (P/LP variants) was established in 29 individuals, and all causal variants were identified by both srGS and lrGS, with no increase in diagnostic yield from lrGS (see Ek et al.Table S2 for additional details). Consistent with prior studies, including ours, lrGS provided additional insights for some variants, by facilitating variant phasing and improved SV resolution. Notably, in one case (RD_P698 in their manuscript), lrGS identified a TR expansion in *FMR1* of 654 repeat units, whereas srGS estimated 76 repeats, below the pathogenic threshold of 200 but within the 55-200 premutation range. This case, along with the two previous diagnostic TRs identified in Cohen et al. and Hiatt et al.,^36,37^ further underscore the utility of lrGS for accurate TR sizing over srGS. While we focused only on those studies that directly reported srGS and lrGS variants, there are many other published studies in which SVs, SNV/indel, or very large CNVs are reported as uniquely discovered by lrGS but would be routinely captured by clinical diagnostic labs using comprehensive srGS analyses. Overall, these studies are consistent with our findings and highlight the current relative incremental improvements in diagnostic yield from lrGS based on existing interpretation criteria when a comprehensive analysis of the srGS is performed.

## DISCUSSION

In this study, we sought to highlight the profound diagnostic contributions of SVs in rare disease diagnostics, and to evaluate the potential value added from lrGS and multiomics among families that remain unresolved by srGS as a front-line strategy. We performed systematic analyses of all types of SVs captured by srGS and identified diagnostic SVs in 5.4% of cases. These results are consistent with our previously published subset of srGS-sequenced families (4.8% diagnostic yield due to SVs).^3^ When considering diagnostic yields, it is critical to consider the genetic architecture of specific phenotypes, as well as to account for pre-screening of cases and families. Here, we note that most of the families included in this study, as part of the Broad CMG or other rare disease research programs, had prior genetic testing that often included a karyotype or microarray, so these results do not reflect the rate of an unscreened rare disease population using srGS as a front-line strategy.^3,83^ These factors introduce ascertainment bias and create challenges when comparing diagnostic yields across studies. One key strength of this study is the systematic, multi-algorithm evaluation and joint calling of SVs from the srGS data. The high resolution and diagnostic potential of comprehensive srGS SV analysis is illustrated by the complexity of variants we were able to resolve: from Alu-mediated compound heterozygous events and disease gene-disrupting inversions to highly complex rearrangements reminiscent of chromothripsis or chromoanasynthesis. These results demonstrate that comprehensive SV analyses are critical to rare disease diagnosis and are a major driver of novel discoveries.

Determining the pathogenicity of SVs has been a long-standing challenge, and the complexity of this task continues to expand with increasing technical capabilities to detect different classes of variants. Current ACMG/ClinGen CNV interpretation standards focus largely on isolated gains and losses, as these are the most unambiguous classes of SVs for prediction of functional impact, whereas balanced and complex SVs present far greater uncertainty in a diagnostics context (as evidenced by our complex mosaic chromoanasynthesis event).^61^ Recent additional criteria have been suggested to evaluate allelic and functional data as well as variants on chrX.^16^ RNA-seq can help to resolve some uncertain cases by providing functional evidence of pathogenicity (ACMG/AMP evidence code PS3). For example, integrating RNA-seq with srGS in BEG_1025, the case of the L1 insertion in *MTM1*, enhanced our ability to classify the SV, reinforcing the value of multiomics approaches in the diagnostic workflow to build evidence towards pathogenicity or benignity of a VUS to reach a definitive classification (P/LP, benign/likely benign).

We identified minimal added diagnostic yield of lrGS in the subset of 96 families assessed here compared to comprehensive srGS analyses. Overall, we identified 15 diagnostic variants in this cohort, with only one case harboring a diagnostic variant that was uniquely observed in the lrGS, representing an incremental added diagnostic yield of 1.04% (1/96). This variant displayed a variable degree of allelic imbalance across multiple srGS libraries (from separate blood draws), as well as allelic imbalance in an lrGS library and in Sanger sequencing on blood and buccal samples. Our results indicate that the allelic distribution in the initial srGS libraries was stochastically lower than the overall mosaic fraction, as evidenced by a repeated srGS sample showing higher (yet still below 50%) allelic fractions of the variant. This diagnostic yield is neither discouraging nor unexpected; novel lrGS diagnostic variant discoveries are at their nascent stages and still reliant on interpretation frameworks dominated by existing technologies, namely ES and srGS. The vast majority of novel variant discovery in lrGS resides in highly repetitive genomic loci such as simple repeats and segmental duplications, where pathogenic variation remains difficult to classify and interpret, yet most of the diagnostic SVs identified in this study (77/79, 98%) are in unique sequences or repeat masked sequences.^24,25,84^ Until lrGS reference datasets expand and metrics related to evolutionary selection are established in such regions of the genome that have not been systematically surveyed, the overall diagnostic increases for patients will continue to be important but modest. Indeed, the most impactful novel discoveries in our cohort resulted from re-analysis of srGS data over time. This included updated analytic pipelines for variant discoveries, novel functional annotations, and continuously emerging reports of novel gene-disease associations and phenotype expansions, such as *RNU4-2* and *GLUL*.^66,85^ These findings are consistent with previous reports on the importance of data reanalysis upon improved methodologies and updated candidate gene lists to increase the number of diagnosis in unsolved families.^86–89^

Given the relatively recent and limited integration of lrGS in the rare disease diagnostic space, the added yield of lrGS remains an open question. Comparing our lrGS findings with previously published cohorts, we identified inconsistencies in how SVs are evaluated in srGS data prior to lrGS, and significant differences in detection capabilities as well as pre-screening prior to introduction of lrGS that will always dramatically influence diagnostic yield.^4,34,90,91^ When we considered rare disease studies that evaluated both srGS and lrGS, the added value of lrGS was largely consistent (0-2%).^36,37,82^ Though this yield is modest at present, the technical advantages of lrGS are manifold.^24,25^ It is clear that lrGS provides transformative capabilities for the discovery of SVs and TRs, as well as novel episignatures.^22,24,84^ Our analyses illustrate both the exciting capabilities of detecting disease episignature matches and other methylation anomalies, as well as the challenge with correlating these results to a causal genetic variant or mechanism. These challenges may suggest limited specificity of methylation analysis, but also reflect limitations of analyzing methylation from blood, including high tissue-specificity of DNA methylation patterns,^92^ cell-type heterogeneity,^93^ lack of reference data for population methylation variation including changes in methylation patterns with age and developmental stage,^94^ and environmental factors, amongst others.^95^ Looking forward, the diagnostic contribution of lrGS will certainly grow as large-scale genome sequencing studies improve our ability to predict and interpret variants in complex regions of the genome, such as tandem repeats and segmental duplications, where the majority of lrGS-exclusive SVs reside.^24,25,96^ Beyond its future potential, lrGS remains a compelling first-pass sequencing approach if the increased cost is acceptable, as it enables the detection of more variants than srGS while providing additional functional information, though analysis is challenged by the lack of reference data to allow filtration of lrGS-exclusive common variants.

Overall, we find that a diverse spectrum of SVs represents a major component of the genetic etiology of rare diseases and must be considered in all unbiased genome sequencing diagnostics. Our analyses project that lrGS offers the potential for a future of expansive analyses of variant classes, episignatures, and phased genomes. However, the added diagnostic yield of lrGS following srGS is likely to be constrained until fundamental advances can be made in establishing the functional consequences of variation in repetitive noncoding sequences of novel repeat-mediated variants in rare diseases. We speculate that transformative advances in ending the diagnostic odyssey for rare disease patients will require a combination of new technologies, dramatic improvements in the annotation and predictive capacity of noncoding variation in the human genome, standardization of analytic frameworks and widespread data sharing from rare disease cohorts. Large rare disease programs such as GREGoR and many others are crucial efforts that are seeking to take these next steps forward to serve the more than half of all rare disease cases that do not harbor easily detectable causal protein coding genetic variants.

## METHODS

### Cohort description

Unsolved individuals with a suspected rare Mendelian disease and their affected and/or unaffected family members were sequenced by the Broad Institute Center for Mendelian Genomics (Broad CMG), a member of the NHGRI GREGoR consortium, recruited through external collaborators typically after prior research exome sequencing was negative (750 families) or through the Rare Genomes Project (712 families) typically after they received an unrevealing result from a clinical diagnostic evaluation. Prior testing varied from no testing to clinical exome. Studies were conducted as part of the NHGRI Centers for Mendelian Genomics^11^ and GREGoR consortium.^39^ Phenotypes were collected as Human Phenotype Ontology (HPO) terms. Approximately half of the families included in this study were previously reported by Wojcik et al.^3^

### Diagnostic status definitions

Families were considered solved if pathogenic/likely pathogenic (P/LP) variant(s) (also defined in this manuscript as diagnostic variants), meeting ACMG/AMP criteria,52 were found in a known disease gene with a moderate or higher level of gene-disease validity evidence, defined by ClinGen’s Gene-Disease Validity curation framework,^97^ and if phenotypes associated with that condition were a clear match for the primary elements of the enrolled proband’s rare disease phenotype. “Probably solved” was used to categorize families meeting solve criteria with one field being imperfect (i.e., variant curated as VUS-high or phenotype match not as strong/specific) (also defined as probably diagnostic variants). ‘Unsolved with a candidate’ and ‘Unsolved’ describes families with insufficient support for variant pathogenicity and/or gene-disease validity for all candidates identified or no candidate was identified respectively. Family solve status in families from external collaborators was determined using slight variations in these criteria, decided by the lead researcher or clinician on that cohort.

### Short-read genome sequencing

#### Sequencing and SNV/indel detection

Short-read GS (srGS) was conducted by the Genomics Platform at the Broad Institute of MIT and Harvard as previously described.^3^ In summary, data were sequenced to achieve average coverage of 30✕ and processed following GATK best practices.^98,99^ Reads were mapped to human genome build 38 (hg38) and SNV/indels were detected using Genome Analysis Toolkit (GATK) HaplotypeCaller. Default filters were applied to SNVs/indels using the GATK Variant Quality Score Recalibration (VQSR) approach. The joint called vcf was loaded into the seqr analysis platform where the annotation pipeline was applied including Variant Effect Predictor (VEP).^65^ Mitochondrial genome (mtDNA) variants were called using the gnomad-mitochondrial pipeline and annotated and filtered as previously reported.^100^

#### Structural variant detection

GATK-SV pipeline was used to detect all classes of SVs, including deletions, duplications, inversions, insertions, translocations, and a spectrum of complex SVs. GATK-SV maximizes sensitivity by harmonizing five algorithms, then adjudicates and re-genotypes SVs from raw read evidence.^18^ GATK-SV is publicly available on GitHub (https://github.com/broadinstitute/gatk-sv) and can be deployed in the google cloud platform via Terra platform. The derived VCF file was annotated with the GATK SVAnnotate tool to predict the functional impact of a given SV, as well as with allele frequencies from population reference databases such as gnomAD SV.^18^

#### TR detection

TRs were genotyped at 71 known disease-associated repeat loci using ExpansionHunter.^101^ The locus specifications, which are publicly available on GitHub (https://github.com/broadinstitute/str-analysis/blob/38dc8bc3805182b07bb63d20fdb76ceae05948ae/str_analysis/variant_catalogs/va riant_catalog_without_offtargets.GRCh38.json), correspond to the same set of loci with population frequencies listed in the gnomAD browser (https://gnomad.broadinstitute.org/short-tandem-repeats?dataset=gnomad_r3). To identify potential pathogenic expansions, we examined individuals with the most expanded genotypes at each locus, comparing them to the pathogenic threshold and gnomAD population frequencies. Genotype quality was further assessed through the review of read visualizations using REViewer.^102^

### Short-read RNA-sequencing

Short-read RNA sequencing (srRNAseq) was performed on a subset of 273 cases as part of this study, of which 9 had a diagnostic SV. RNA from whole blood, cultured fibroblasts, or muscle were sequenced using a stranded, polyA-tailed kit (Illumina TruSeq Stranded), generating 50-150 million 150 bp paired-end reads per sample (increased read depth for more recently sequenced samples). Sequencing data was processed using a pipeline adapted from GTEx (GTEx Consortium 2020).^103^ Briefly, FASTQ files were aligned to the GRCh38 reference sequence using STAR-2.7.10b in 2-pass mode, and duplicates were marked with Picard. Expression quantification was performed with RNA-SeQC v2.4.2^104^ to generate transcripts per million (TPM) read counts. Quality control metrics were produced using FastQC v0.12.1 and RNA-SeQC, then summarized with MultiQC.^105^ OUTRIDER^106^ was applied to normalized TPM read counts for gene expression outlier detection, and FRASER v1.99^107^ was run on aligned BAMs to detect splicing outliers, using 104 whole blood, 199 cultured fibroblast, or 384 muscle RNA-seq samples from our own cohort of individuals with rare disease. Volcano plots were generated from both tools. For FRASER’s filterExpressionAndVariability step, the minimum read count in at least one sample was set to 2 and minimum ΔѰ to 0.05.

### Long-read genome sequencing

#### Sample selection

For a subset of participants (242 individuals, 96 families), lrGS data were generated using Oxford Nanopore Technology (ONT) (61 individuals from 20 families, previously reported by Negi et al., 2025)^42^ or Pacific Biosciences (PacBio) (181 individuals from 76 families, inclusive of one quintet, one quartet, 46 trios, 6 duos, and 22 singletons). Families in the PacBio cohort were selected if meeting one of the following criteria: i) families with neurodevelopmental phenotypes and samples from both unaffected parents available for sequencing (n=47 families, one of the parents failed sequencing), ii) families with a single P/LP variant or rare VUS in a recessive disease-associated gene with phenotypic overlap, where lrGS could assess for a second hit (n=10), and iii) families with a clinical diagnosis, where a targeted gene-list would focus the analysis efforts (e.g., Hereditary Hemorrhagic Telangiectasia [HHT], Limb Girdle Muscular Dystrophy [LGMD]) (n=19). PacBio sequencing of these families has not been reported previously. Not all samples had SV analysis completed at the time of cohort selection: 40/76 Pacbio families and 5/20 ONT families had not had SV analysis from the srGS data. To account for this, families with diagnostic variants detectable by srGS are excluded when calculating the added diagnostic yield of lrGS.

#### Sequencing

Briefly, HMW DNA was extracted from whole blood and sheared, resulting in a peak of approximately 50 kb. Sequencing was performed on the PromethION 48 sequencer using R10.4.1 flow cells. For PacBio CCS library preparation, high molecular mass genomic DNA was first purified using the following extraction kits over the course of the project: QIAsymphony DSP DNA Midi Kit (96), Promega Maxwell® HT 96 gDNA Blood Isolation System (A2671), Qiagen Magattract HMW Kit and PacBio Nanobind PanDNA Kit. At least 4 μg of high molecular weight genomic DNA (> 50% of fragments ≥ 40 kb) was sheared to ∼15 kb using the Megaruptor 3 (B06010003; Diagenode), followed by DNA repair and ligation of PacBio adapters using the SMRTbell Prep Kit 3.0 (102-141-700). Each library was subsequently size-selected for 10 kb ± 20% using the PippinHT with 0.75% agarose cassettes (Sage Science). After quantification with the Lunatic (Unchained Labs), libraries were diluted to 250 pM per single molecule, real-time (SMRT) cell, hybridized with PacBio standard sequencing primer, and bound with SMRT sequencing polymerase using the Revio polymerase kit (102-739-100). CCS sequencing was performed on the Revio instrument using 25M SMRT Cells (102-202-200) and Revio Sequencing Plate (102-587-400), with a 2-hour pre-extension time and 24-hour movie time per SMRT cell. Quality filtering, basecalling, and adapter marking were done automatically on board the Revio. Error correction for reads generated in CCS mode was performed on-board the PacBio Revio with the vendor’s ccs software^108^ and settings --all --subread-fallback --num-threads 232 --streamed <movie_name>.consensusreadset.xml --bam <movie_name>.reads.bam. With these settings, all reads from the instrument (including those failing error correction) are presented in a single BAM file for downstream analysis.

#### Data processing and SNV/indel detection

ONT and PacBio lrGS data were aligned to the GRCh38 reference genome with Minimap2 version 2.24.^109^ Small variants (SNV/indels) were called with DeepVariant v1.3.0/v1.8.0^110^ and joint-genotyped with GLNexus.^111^ The joint-called VCF files were functionally annotated with the Ensembl Variant Effect Predictor (VEP, release 113),^65^ precomputed SpliceAI scores,^112^ gnomAD v4.1 allele frequencies,^113^ and classification plus review status in ClinVar.^114^ In combination with the SVs, the small variants were filtered using custom scripts to recapitulate “*de novo*/dominant” and “recessive” seqr searches with both “restrictive” and “permissive” thresholds for potential pathogenicity (Table S1, see Variant filtering and interpretation).

#### Structural variant detection

For ONT samples, SVs were detected as described.^42^ Briefly, reads were assembled using Shasta and Hapdup, and SVs were detected from the resulting diploid assembly relative to the GRCh38 reference using hapdiff. In parallel, reads were aligned to GRCh38 using Minimap2 and SVs were detected using Sniffles. For PB samples, SVs were detected from aligned reads using Sniffles v2.0.6, pbsv v2.9.0, and HiFiCNV v0.1.7. For each technology cohort, SVs were merged at the intrasample level using truvari collapse^115^ with settings “--intra -k first --refdist 500 –pctseq 0.90 --pctsize 0.90” and were then merged at the cohort level using truvari collapse with settings “”-k first --refdist 500 --pctseq 0.90 --pctsize 0.90”. Functional annotations were added using SVAnnotate from GATK-SV^18,116^ using the MANE Select v1.2 gene dataset. Due to the tendency of Sniffles, pbsv, and hapdiff to classify duplications as insertions, we annotated insertion calls with the functional impact that they would have as either insertions or duplications. Each variant was further annotated with allele frequencies from the following cohorts: gnomAD SV v4.1.0,^18^ 100 samples from the 1KGP-ONT Consortium,^117^ and lrGS from Genomic Answers for Kids (GA4K) (n=726).^37^ Additionally allele frequencies of SVs for these samples identified in the srGS were annotated. Variants were matched to SVs from these external callsets using code adapted from GATK-SV.^18^ lrGS SV data were analyzed outside of seqr. SNVs/indels and SV were filtered according to standard seqr searches (Table S1, see Variant filtering and interpretation).

#### Rearrangement breakpoint detection

To detect rare *de novo* rearrangement breakpoints, BigClipper^26^ was run on all lrGS affected samples with parameters “--max_unique_breakends 10 --min_cluster_count 5 --min_dist 20000” and on all unaffected samples with parameters “--max_unique_breakends 50 --min_cluster_count 1 --min_dist 5000.” Output VCFs were merged using bcftools merge with parameters “-m none” and clustered using bedtools cluster “-d 50”. Clustered breakpoints found in more than one sample were removed, and breakpoints from affected samples were manually evaluated. A median of four candidates were detected per affected sample.

#### TR detection and analysis

TRGT v1.5.1^69^ was used to genotype 71 known disease-associated TR loci, listed in https://github.com/broadinstitute/str-analysis/blob/25c6f87f9729674137a08b31285b2a25d034e80c/str_analysis/variant_catalogs/cat alog.GRCh38.TRGT.bed. Analysis was performed by using a custom script https://github.com/broadinstitute/str-analysis/blob/25c6f87/str_analysis/check_combined_results_tsv_for_pathogenic_repeats.py to sort the results at each locus by allele size and display them for manual review while taking into account the locus-specific inheritance modes and pathogenic thresholds listed in https://github.com/broadinstitute/str-analysis/blob/25c6f87/str_analysis/variant_catalogs/variant_catalog_without_offtargets.GRCh38.json. For expansions above the pathogenic threshold, read visualizations were generated using the TRGT plot functionality and then manually reviewed for genotype quality, interruption patterns, and motif composition using the flipbook tool.^102^

#### Paraphase analysis

The PacBio tool Paraphase (https://github.com/PacificBiosciences/paraphase) v3 was applied to all lrGS samples. Fourteen clinically-relevant regions (*SMN1/SMN2, RCCX* module*, PMS2, STRC, IKBKG, NCF1, NEB, F8, CFC1, OPN1LW/OPN1MW, HBA1/HBA2, GBA, CYP11B1/CYP11B2, CFH* cluster) were analyzed from the output JSON, vcf, and realigned BAM files.

#### Methylation

Site-specific methylation levels were compiled from aligned reads using modkit (ONT samples) or pb-CpG-tools (PacBio samples). The respective output files were converted to bismark format and processed using the BSmooth^71^ package to detect differentially methylated regions (DMRs) as follows. Data was first smoothed using the BSmooth function. For each affected sample, the smoothed data was filtered to retain sites where the affected sample and at least two unrelated samples had at least five total reads. Differentially methylated CpG sites were identified using BSmooth.tstat, comparing the affected sample to all unrelated samples and estimating variance among the latter group. These sites were clustered into differentially methylated regions (DMRs) using BSmooth.dmrFinder, using a t-stat cut-off corresponding to a significance level of p<0.05 with Bonferroni correction for the number of CpG islands or candidate cis-regulatory regions (cCREs) from ENCODE Registry V3^72^ in the genome multiplied by the number of samples being evaluated. Candidate DMRs were filtered as follows: 1) DMRs had to overlap either CpG islands or cCREs by a minimum of 80%, and 2) DMRs had to contain at least five CpGs, a t-stat standard deviation lower than 0.1, and a minimum absolute value of 0.3 mean methylation difference between the affected sample and the control group. These candidate DMRs were filtered for association with known disease-linked genes via overlap with at least one of the following categories: 1) activity-by-contact (ABC)-derived enhancers^118^ associated with OMIM^119^ genes in any tissue type (minimum 10% overlap), 2) promoters (defined as 1 kb upstream of the transcription start site) of OMIM genes (minimum 10% overlap), and/or 3) overlap with the full gene body of OMIM genes using MANE^120^ v1.2 transcript annotations (any amount of overlap).

#### Imprinting disorders

Methylation at 14 known imprinting disorder loci was inspected by running NanoImprint,^70^ and outputs were manually inspected for each sample to identify potential imprinting irregularities, with follow-up inspection of read alignment and phasing performed in IGV. Poor read alignment at the H19 locus often resulted in very low coverage and uninformative imprinting results, as previously reported when using the GRCh38 reference,^70^ and as a result it was excluded from analysis.

#### Episignatures

Sample episignatures were matched to reported disease episignatures using NSBEpi.^74^ Briefly, for each of 34 previously reported episignatures,^44^ a support vector machine (SVM) classifier was trained using the data released in that study, representing one case example of methylation levels at the episignature-relevant CpG sites and 34 examples of methylation levels in the absence of the disorder. The classifier was then applied to the methylation levels of each sample in our study at the episignature-relevant CpG sites, with confidence scores over 0.3 being considered positive matches to the disorder episignature.^74^ Additionally, episignatures for 16p11.2 deletions and CHD8 variants were obtained from Siu et al.^73^ and analyzed using the same approach.

### Long-read RNA sequencing

#### Sample selection and Kinnex Library Preparation

We selected 20 probands and their parents with previous PacBio lrGS for lrRNAseq (n=60). Inclusion criteria were sufficient RNA quantity for Kinnex library preparation. Trios were prioritized based on higher RNA quality scores (RQS). An aliquot of 300ng of total blood RNA is used as the input for Kinnex™ full length cDNA synthesis and amplification (PacBio Iso-Seq express 2.0 kit, 103-071-500). The amplified cDNA is then quantified by Qubit and Tapestation (Qubit dsDNA HiSens, QUBDSDNA500KT, and HiSense D5000 ScreenTape, 50675592) and barcoded cDNA was then pooled before proceeding to 8-fold Kinnex PCR (PacBio Kinnex PCR 8-fold kit, 103-071-600). Samples then were programmatically concatenated into single molecules optimal for long-read sequencing (Kinnex full-length RNA kit, 103-072-000). Success of array formation was evaluated through qubit quantification and Agilent TapeStation fragment size QC (Qubit dsDNA HiSens, QUBDSDNA500KT, and GenomicDNAScreenTape,50675366).

#### Pacific Biosciences Revio Sequencing

Each pooled Kinnex library undergoes sequencing preparation using the Revio polymerase kit (Pacific Biosciences, 102-739-100). Using the Sample Setup page in SMRTLink, the appropriate volumes of each reagent are calculated by factoring in the insert size, sample concentration, and target plate loading concentration of 150pM. The run design is set up with the Application Type “Kinnex full-length RNA” and Library Type “Kinnex.” Samples are loaded onto the sequencer within 24 hours of completed sequencing preparation and are sequenced using the 24 hour movie setting. After sequencing and post processing, which occurs on the instrument, the data is transferred to a Google Cloud bucket for downstream analysis.

#### Analysis of lrRNAseq data

SQANTI3^121^ was used to map each assembled transcriptome to the human reference (Gencode v48, hg38) using the default settings in the sqanti3_qc.py script. To subselect on filtered transcripts, the rules and random-forest filters were identified, using the default parameters for both in the sqanti3_filter.py script (Supplementary Figure S9). For each gene and variant proposed, we chose several metrics to determine if further manual review was necessary. 1) The TPM of the proband vs the parents. 2) The proportion of gene expression coming from non-FSM transcripts in the proband versus the parents, before and after filtering. 3) Whether any novel splice acceptor or donor detected in the proband overlapped with a rare or *de novo* variant. 4) Whether there was a fusion transcript detected for the proband in that gene.

We generated several lists for review of genes associated with altered transcriptomes, with filters being chosen to limit the number of genes chosen to review. First, we selected genes containing Novel-In-Catalog and Novel-Not-in-Catalog transcripts that passed both the rules and ML filters and were >10% of the TPM of the total gene’s TPM, provided that the gene also contained either a *de novo* variant or rare LOF variant. We also identified any gene containing a rare pLoF variant where a different rare variant lay within 20 bp of a novel splice acceptor or donor. No *de novo* variants were within 20bp of a novel splice donor or acceptor. Finally, we identified any gene with a mapped fusion transcript that also contains either a *de novo* variant or a rare pLoF variant. For gene lists related to *de novo* variants, we also excluded any transcript structure that was present in either parent.

#### Variant filtering and interpretation

SVs in the srGS data were analyzed by Broad RGP variant analysts in the open-source, web-based genomic analysis platform, seqr (v1.0-ab24c2bc). Predefined family-based searches for “*de novo*/dominant” and “recessive” variants with both “restrictive” and “permissive” thresholds for potential pathogenicity were applied as previously described.^122^ Briefly, for SV “Restrictive” searches, high quality, rare variants with moderate-high functional impact are returned, including LoF, intragenic exon duplication, and copy-gain variants. For SV “Permissive” searches, additional lower impact variants are considered, this includes partial exon duplications, partial gene duplications, intronic SVs, inversion spans, UTR, promoter, and breakend exonic SVs.

SVs returned by each seqr search were carefully reviewed for clinical relevance to the proband’s phenotype and visually inspected using the Integrated Genomics Viewer (IGV v.2.18.2). For variants in genes with a reported Mendelian disease association (based upon the OMIM database^119^ and a PubMed literature search), the proband’s phenotype and the inheritance pattern were assessed for consistency with reported cases and for potential novel disease mechanisms (i.e., alternative modes of inheritance). For variants in genes not yet associated with Mendelian disease, we mostly focused on dosage sensitive genes, defined as genes that are LoF intolerant (LOEUF < 0.3 and/or pLI ≥ 0.9), demonstrate haploinsufficiency (i.e., deletion intolerance, pHaplo > 0.86) or triplosensitivity (i.e., duplication intolerance, pTriplo > 0.94).^18,113,123^ We cross-referenced external databases to collect evidence supporting a potential role in disease, for example, tissue RNA and protein expression pattern (GTEx, Human Protein Atlas),^124,125^ phenotype overlap with animal models (MGI, IMPC, ZFIN),^126–128^ and review of available literature on the function of the encoded protein (PubMed, GeneCards).^129^

lrGS data were analyzed external to seqr, using custom scripts to recapitulate the “*de novo*/dominant” and “recessive”, “restrictive” and “permissive” family-based filtering strategies applied in seqr for the srGS, encompassing the analysis of both SNV/indel and SV variants as reported in Table S1. Briefly, for “Restrictive” searches, high quality, rare variants with moderate-high functional impact are returned. For SVs, LoF, intragenic exon duplication, and copy-gain variants are returned. For SNVs/indels, LoF, missense, and in-frame indel variants are returned, as well as variants predicted to impact splicing by SpliceAI ≥0.2.^112^ For “Permissive” searches, additional lower impact variants are considered. For SVs, this includes partial exon duplications, partial gene duplications, intronic SVs, inversion spans, UTR, promoter, and breakend exonic SVs. For SNV/indels, synonymous, splice region, UTR, and regulatory variants are returned, as well as variants predicted to impact splicing by SpliceAI ≥0.1. Permissive searches also have reduced stringency on call quality (GQ ≥ 30, AB ≥ 0.1). These searches incorporate both SVs and small variants (SNV/indels), to allow for the detection of compound heterozygous diagnoses involving both variant types. In addition, for families with no candidate variant detected by our analysis pipeline and either: i) a strong suspicion of a specific genetic diagnosis based on phenotype (e.g., ENG and ACVRL1 for probands with hereditary haemorrhagic telangiectasia) or ii) a confident single hit in a recessive disease gene, the aligned sequencing reads were visually inspected using the Integrated Genomics Viewer (IGV v.2.18.2) for missed variants and deviant patterns in read mapping or methylation.

Variants were classified according to the ACMG/AMP criteria.^60^ Pathogenic/Likely Pathogenic (P/LP) as well as variants of uncertain significance (VUS) leaning towards LP (VUS-high) were sent for clinical confirmation and returned to the respective families via their local clinician for RGP families and handled according to their IRB protocols by external collaborators. Only P/LP variants were considered to be definite diagnoses, VUS-high were considered to be probable diagnoses.

#### Literature searches to compare diagnostic yield

We systematically reviewed all articles returned by PubMed searches for “long-read sequencing rare disease” and “long-read sequencing neurodevelopmental” available at the time of this study. Out of 464 publications (search date Feb 25th 2026), only three studies: Cohen et al. (2022),^37^ Hiatt et al. (2024),^36^ and EK et al. (2026),^82^ performed both srGS and lrGS on more than 50 rare disease cases and clearly distinguished the variants identified by each sequencing technology.

## DATA AVAILABILITY

Genomic and phenotypic data from the Broad CMG is available via dbGaP accession numbers phs003047 (GREGoR) and phs001272 (CMG). Access is managed by a data access committee designated by dbGaP and is based on intended use of the requester and allowed use of the data submitter as defined by consent codes. Code used in this study is available at https://github.com/talkowski-lab/sv-rare-diseases.

## ETHICAL CONSIDERATIONS

All participants in the Rare Genomes Project were consented on a protocol approved by Mass General Brigham IRB protocol 2016P001422) and families recruited by external collaborators through local IRB protocols with coded samples and metadata shared for secondary use with the Broad CMG (Mass General Brigham IRB protocol 2013P001477).

## ACKNOWLEDGMENTS

This work was supported by the NHGRI GREGoR Program (U01HG011755, U24HG011746), NICHD (R01HD081256, R01HD105266), NIMH (R01MH115957). Reagents and/or research support was provided by Pacific Biosciences, Oxford Nanopore Technologies, and Illumina Inc. Sequencing and analysis of additional Broad CMG cohorts were funded by the NHGRI grants UM1HG008900 (with additional support from the National Eye Institute, and the National Heart, Lung and Blood Institute), and R01HG009141, NIDCR (R01DE031261, R01DE030342, R00DE026824), National Institute of Diabetes and Digestive and Kidney Diseases (NIDDK) RC2DK122533, National Eye Institute (RO1EY012910), the Simons Foundation Autism Research Initiative (SFARI #19405), and in part by the Chan Zuckerberg Initiative Donor-Advised Fund at the Silicon Valley Community Foundation (funder DOI 10.13039/100014989) grants 2019-199278, 2020-224274, 2022-316726, 2022-247486 (https://doi.org/10.37921/236582yuakxy). Subject ascertainment, enrollment, and data collection for MAN_* and BEG_* cases was made possible by support from the Gene Discovery Core of the Manton Center for Orphan Disease Research, and MDA1457288 (https://doi.org/10.55762/MDA.1457288.pc.gr.235448) from the Muscular Dystrophy Association, USA, Cure ADSSL1 and Cure CMD. Confirmatory Sanger sequencing of variants in these cases was performed by the Boston Children’s Hospital IDDRC Molecular Genetics Core Facility funded by P50HD105351 from the NICHD, and as part of the Boston Children’s Hospital Children’s Rare Disease Collaborative study. A.S-J. was supported by the Mass General Brigham T32 training grant fellowship in the Center for Genomic Medicine (2T32HG010464-06). S.L.S. and L.E.C were supported by fellowships from the Manton Center for Orphan Disease Research at Boston Children’s Hospital. V.S.G. was supported by NIH/NHGRI grant K23AR083505 and the BroadIgnite Award. O.M. is supported by the Hazem Ben-Gacem Tunisia Medical Fellowship Fund. KÕ is supported by the Estonian Research Council Grants PRG471 and PRG2040. S.P. is supported by the Estonian Research Council Grant PSG774. J.A.J. was supported by T32GM007748, 5T32NS007473, 5T32EY007145, and the Harvard Medical School William Randolph Hearst Fund. E.C.E. is a Howard Hughes Medical Institute Investigator. MYO-SEQ was funded by Sanofi Genzyme, Ultragenyx, LGMD2I Research Fund, Samantha J. Brazzo Foundation, LGMD2D Foundation and Kurt+Peter Foundation, Muscular Dystrophy UK, and Coalition to Cure Calpain 3. The Rare Disease Flagship acknowledges financial support from the Royal Children’s Hospital Foundation, the Murdoch Children’s Research Institute, The Harbig Foundation, The Fox Family Foundation, The Andrew and Geraldine Buxton Foundation and The Pierce Armstrong Foundation. The research conducted at the Murdoch Children’s Research Institute was supported by the Victorian Government’s Operational Infrastructure Support Program. This research was supported in part by the Intramural Research Program of the National Institutes of Health (NIH), the National Eye Institute (EY000564). The contributions of the NIH author(s) were made as part of their official duties as NIH federal employees, are in compliance with agency policy requirements, and are considered Works of the United States Government. However, the findings and conclusions presented in this paper are those of the author(s) and do not necessarily reflect the views of the NIH or the U.S. Department of Health and Human Services.

