## Supplemental Materials for "Structural variant discovery and diagnostic impact in rare diseases from short-read and long-read sequencing"

### Supplemental Data

#### Improved coverage of clinically relevant regions by long-read sequencing

Using GRCh38-aligned reads with a mapping quality (MQ) of ≥10 and depth (DP) of ≥10, long-read genome sequencing (lrGS) demonstrated improved coverage of genes over short-read genome sequencing (srGS), in particular clinically significant genes reported as challenging to sequence due to repetitiveness or high sequence homology,[1,2] such as *CBS, CRYAA, CYP21A2, KCNE1,* and *SMN1* (Figure 3A). Calculating the median gene-length covered per gene across all samples, 260 protein-coding genes and 11 disease-associated genes had ≥90% of their entire length covered by lrGS only (Figure 3B). Focusing on exons of protein-coding genes, where P/LP variants associated with Mendelian disease are most often located,[3] 309 genes had at least one exon well covered by lrGS only, of which 20 were disease-associated genes. These disease-associated genes included 4 genes (*GRAP, NSF, SIK1,* and *TRAPPC10*) that have not previously been reported among the challenging clinical significant genes by Mandelker et al., and Wagner et al., and include two dominant NDD-associated genes, NSF and SIK1 (Figure 3C). Limiting to the genomic positions of 66,249 variants already reported as P/LP in ClinVar with at least a 2-star review status (criteria provided, multiple submitters, no conflicts),[3] 39 variant positions were well covered by lrGS only, all within 5 genes (*CBS, CRYAA, IKBKG, NEB,* and *SMN1*). The increased number of genes, exons, and P/LP variant positions well covered by lrGS indicate added value for Mendelian disease diagnosis.

#### Genome coverage analysis

srGS and lrGS data aligned to the hg38 reference genome were analyzed using mosdepth[4] to produce a per-sample bed file of genomic positions where reads were aligned at MQ ≥10 and DP ≥10. Gene annotation, including gene type (e.g., protein-coding), start position, stop position, and exon positions were annotated using Ensembl Biomart.[5] For exon analyses, the MANE Select transcript was used for annotation.[6] Genomic features (genes, exons, positions of ClinVar P/LP variants) were considered well covered when across all samples, a median of ≥90% of the length was covered at MQ ≥10 and DP ≥10. For positions on chrX, only samples from female individuals were considered. ClinVar 2-star P/LP variants were downloaded from the ClinVar (https://ftp.ncbi.nlm.nih.gov/pub/clinvar/, last accessed 01.09.2025)[7] and filtered to variants within disease-associated genes reported in the OMIM database (last accessed 14.01.2025).[8]

### Supplemental Figures

**Figure S1.** Structural variant (SV) diagnostic rate for 1,462 families undiagnosed prior to srGS analysis. **A)** By primary phenotype (indication for genetic testing). Only phenotypes with ≥10 families were included in this analysis. **B)** By availability of parental data for variant phasing. Proportions of families are displayed on the y-axis with the total counts of families displayed above the bars. Diagnosed with SV includes compound heterozygous SV plus small variant (SNV+indel) diagnoses.

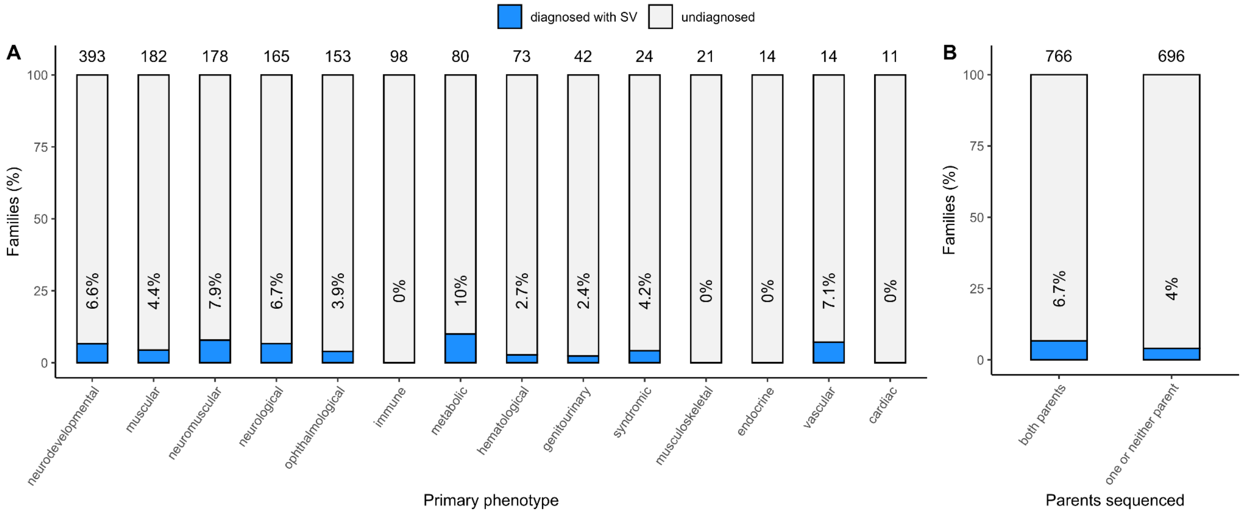

**Figure S2. Validation of TR expansion in NOTCH2NLC locus.** Fluorescence amplicon length analysis revealed that the size of normal repeat (blue arrow) was 32 and the size of abnormal repeat (red arrow) was ~145 repeat (although the peak of the PCR product was widely distributed).

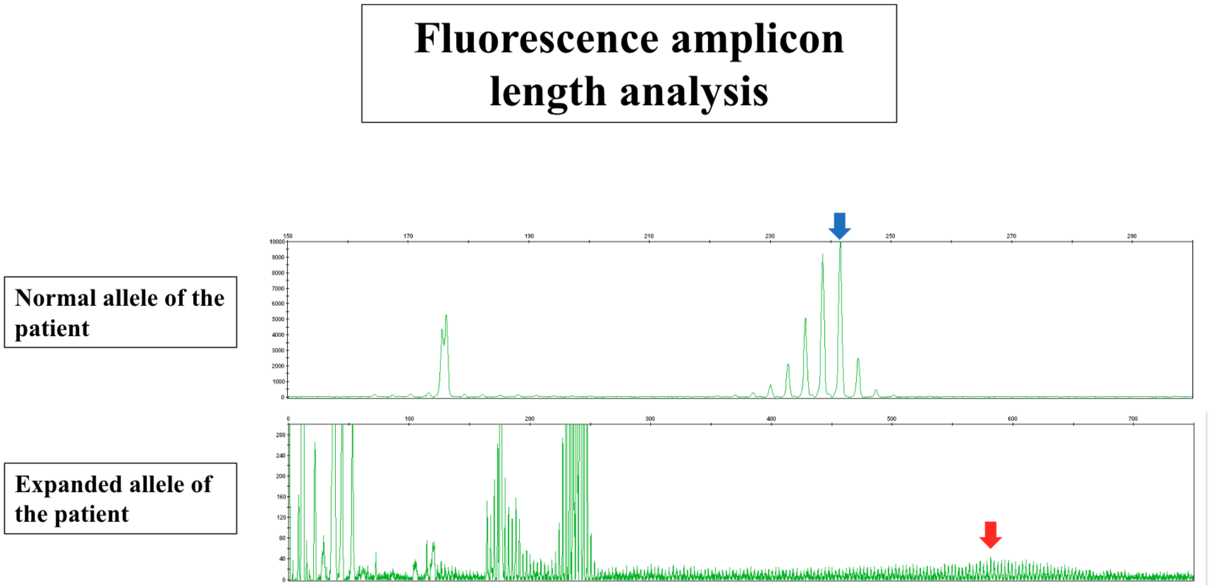

**Figure S3. Complex structural variant (SV) identified in the EA family. A)** Schematic of the complex SV. A 116 kb duplication of chr8 (chr8:65,164,805-65,281,089) is inserted into intron 43 of DMD. A 126 bp segment of intron 43 (chrX:32,258,778-32,258,903) is duplicated and flanks the chr8 insertion on both sides. An additional 13 bp insertion (insGCCTTTGCCCACA) is present adjacent to the 126 bp duplication (not shown). **B)** srGS read alignments supporting the chr8 duplication. **C)** srGS read alignments at the insertion site in intron 43 of DMD. 13/24 reads at the breakpoint support the insertion in this male individual, suggesting somatic mosaicism. **D)** Sashimi plot showing aberrant splicing in the proband (top) relative to GTEx skeletal muscle (bottom). A novel splice junction is supported by 6 reads, while 3 reads retain the wild-type junction, consistent with the SV being in mosaicism state. Abnormal retention of intron 40 is also observed. **E)** srRNAseq (from muscle tissue) read alignments in the EA family. The chromosomal fusion between chrX and chr8 generates multiple novel splice junctions. Split reads map from the splice donor of DMD exon 43 to three distinct acceptor sites on chr8 (BLAT alignment results, left panel).

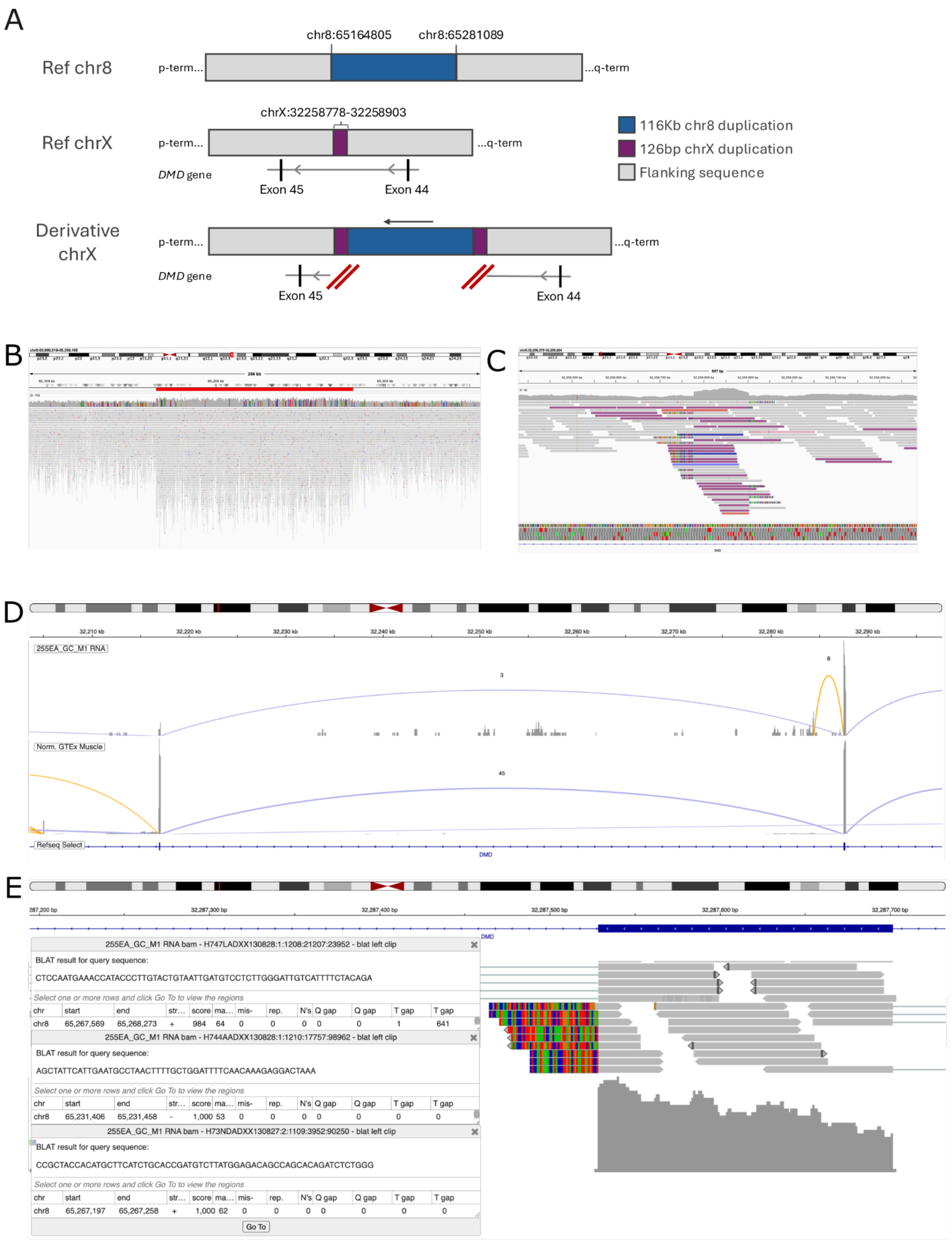
A
chr8:65164805
chr8:65281089
Ref chr8
p-term...
...q-term
chrX:32258778-32258903
Ref chrX
DMD gene
Exon 45
Exon 44
116Kb chr8 duplication
126bp chrX duplication
Flanking sequence
Derivative chrX
B
C
D
255EA_GC_M1 RNA
Norm. GTEx Muscle
Refseq Select
DMD
E
255EA_GC_M1 RNA bam - H747LADXX130828:1:1208:21207:23952 - blat left clip
BLAT result for query sequence:
CTCCAATGAAACCATACCCTTGTACTGTAATTGATGTCCTCTTGGGATTGTCATTTTCTACAGA
Select one or more rows and click Go To to view the regions
chr start end str… score ma… mis- rep. N's Q gap Q gap T gap T gap
chr8 65,267,569 65,268,273 + 984 64 0 0 0 0 0 1 641
255EA_GC_M1 RNA bam - H744AADXX130828:1:1210:17757:98962 - blat left clip
BLAT result for query sequence:
AGCTATTCATTGAATGCCTAACTTTTGCTGGATTTTCAACAAAGAGGACTAAA
Select one or more rows and click Go To to view the regions
chr start end str… score ma… mis- rep. N's Q gap Q gap T gap T gap
chr8 65,231,406 65,231,458 - 1,000 53 0 0 0 0 0 0 0
255EA_GC_M1 RNA bam - H73NDADXX130827:2:1109:3952:90250 - blat left clip
BLAT result for query sequence:
CCGCTACCACATGCTTCATCTGCACCGATGTCTTATGGAGACAGCCAGCACAGATCTCTGGG
Select one or more rows and click Go To to view the regions
chr start end str… score ma… mis- rep. N's Q gap Q gap T gap T gap
chr8 65,267,197 65,267,258 + 1,000 62 0 0 0 0 0 0 0
Go To

**Figure S4. Sanger sequencing validation of the de novo SNV variant in CASK identified in RGP_1777_3 proband** in blood sample **A)** and saliva **B)**. The variant is supported to be mosaic, at a 20-35% in the Sanger trace taking in account the variability of signals from the dyes.

**A)**

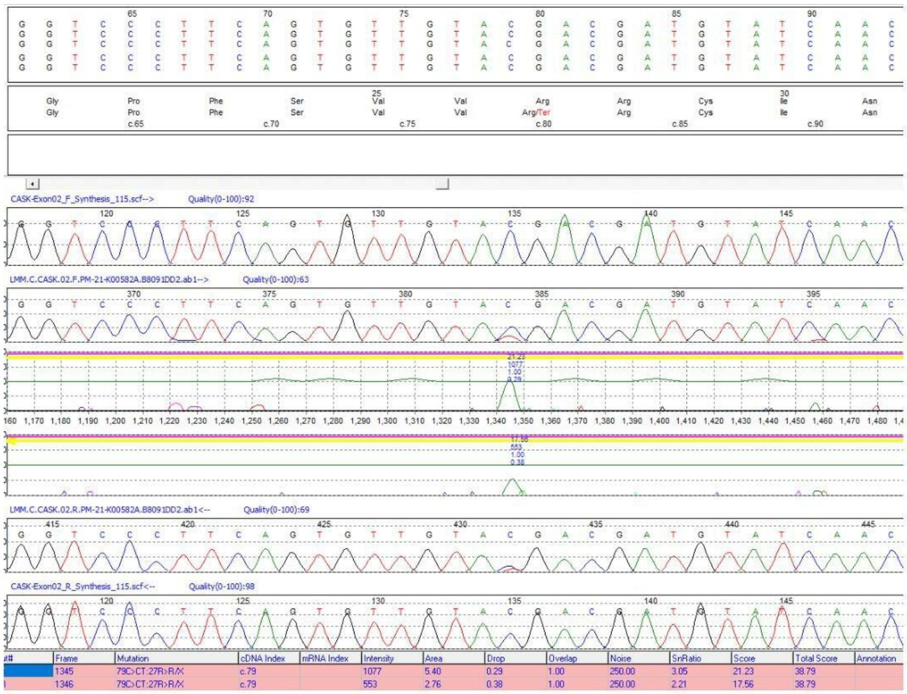


**B)**

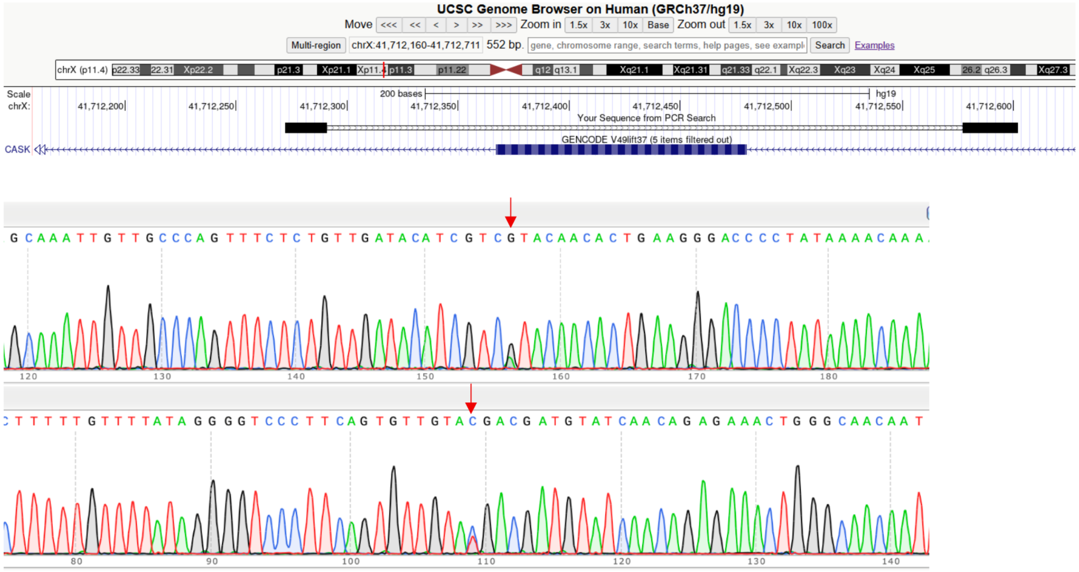

**Figure S5. Breakpoint resolution of SVs with lrGS.** Each panel shows a view from the Integrative Genomics Viewer for a diagnostic SV. The top row shows short reads, followed by long reads, long-read-based assembly, and a relevant repeat track (either LINE, SINE, or segmental duplication (SD), depending on which element was present at the breakpoint). **A)** 1.11 Mb deletion in sample RGP_2210_3 spanning multiple genes including CNOT2 (genes not shown), with breakpoints in two LINE elements. **B)** Mosaic deletion in CDKL5 in sample RGP_2158_3, with breakpoints in Alu elements. **C)** Homozygous deletions in CYB5R3 in samples RGP_2131_3 and RGP_2131_4, with breakpoints in Alu elements**. D)** Tandem duplication in ENG in sample RGP_1890_3, with breakpoints in Alu elements. **E)** Deletion in HECTD4 in sample RGP_1830_3, with breakpoints in Alu elements. **F)** 1.55 Mb deletion at 2q11.2 (spanning multiple genes, not shown) with breakpoints in segmental duplications.

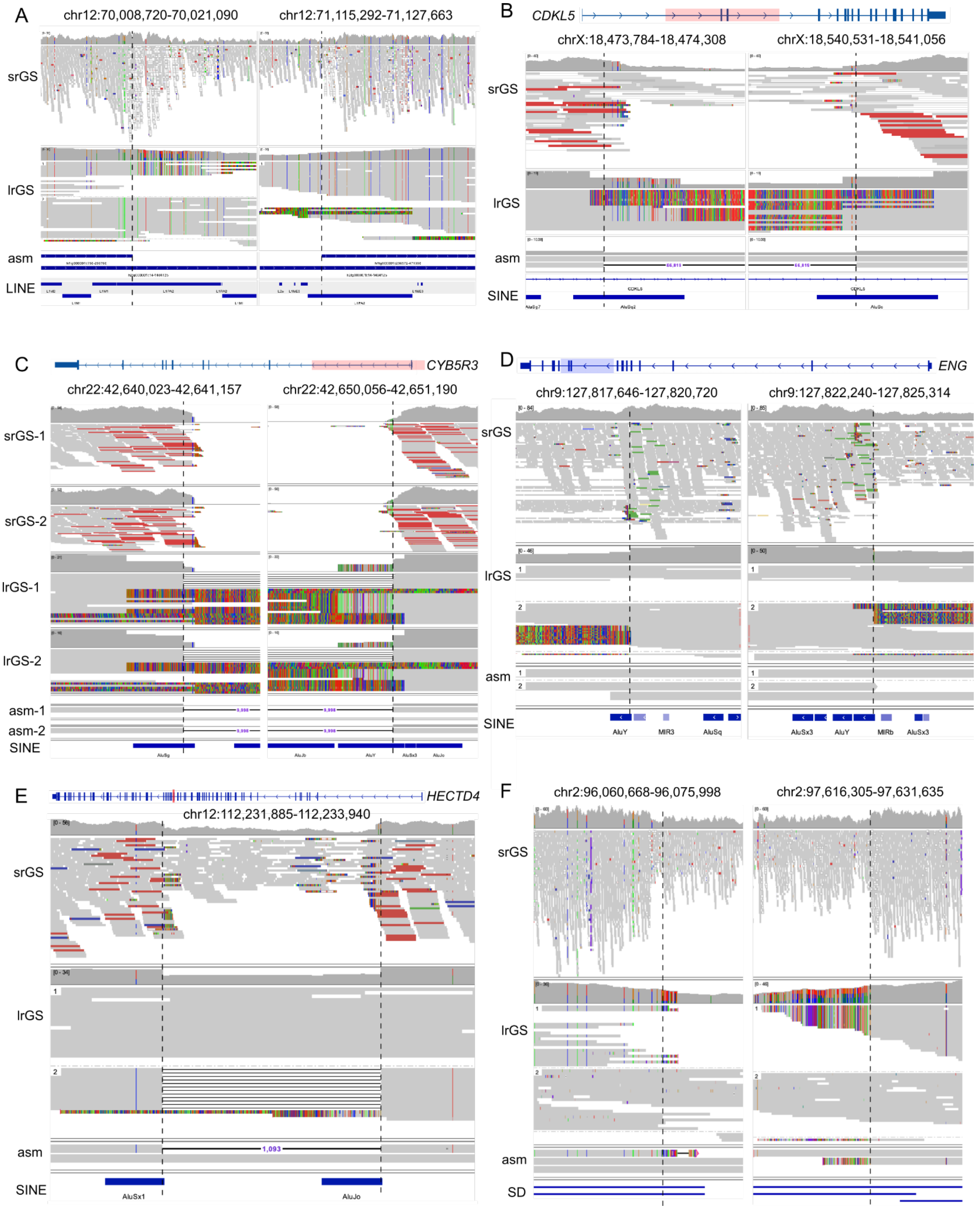

A
chr12:70,008,720-70,021,090
chr12:71,115,292-71,127,663
srGS
lrGS
asm
LINE
B
CDKL5
chrX:18,473,784-18,474,308
chrX:18,540,531-18,541,056
srGS
lrGS
asm
SINE
C
CYB5R3
chr22:42,640,023-42,641,157
chr22:42,650,056-42,651,190
srGS-1
srGS-2
lrGS-1
lrGS-2
asm-1
asm-2
SINE
D
ENG
chr9:127,817,646-127,820,720
chr9:127,822,240-127,825,314
srGS
lrGS
asm
SINE
AluY
MIR3
AluSq
AluSx3
AluY
MIRb
AluSx3
E
HECTD4
chr12:112,231,885-112,233,940
srGS
lrGS
asm
1,093
SINE
AluSx1
AluJo
F
chr2:96,060,668-96,075,998
chr2:97,616,305-97,631,635
srGS
lrGS
asm
SD

**Figure S6. Added value of lrGS for sequence variation. A)** Compound heterozygous variants in HECTD4 (RGP_1830) including (1) an inherited maternal (“hap1”) SNV at a splice donor site and (2) a de novo 1.06 kb deletion spanning an exon on the paternal haplotype (“hap2”). Variants were confirmed to be in trans using read-backed phasing with lrGS without the need for parental data. **B)** Mosaic de novo chromoanasynthesis case (RGP_1316) involving at least 24 breakpoints between 10 chromosomes and at least 9 duplicated regions. Outer circle, chromosome coordinates in Mb units. Middle circle, coverage track derived from srGS data averaged in 1 kb windows, with duplications shown in blue. Inner lines denote breakpoints, with intrachromosomal connections in green and interchromosomal connections in black (detailed breakpoint information in Table S4).

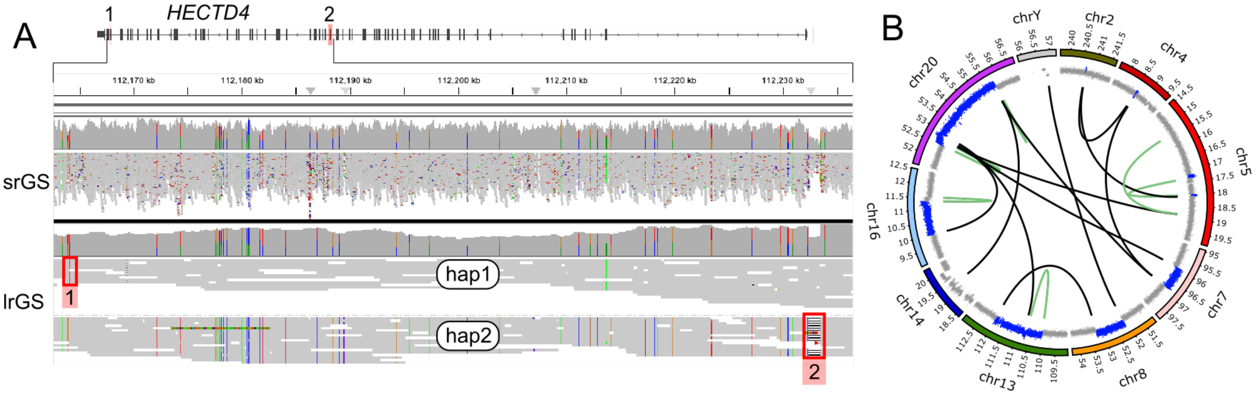

**Figure S7. Genome coverage by srGS and lrGS. A)** Median gene length (%) covered at MQ ≥10 and DP ≥10 across samples, for genes within each gene group (left y-axis, colored points). Genes (%) in the group that are well covered (defined as median gene length covered ≥90% at MQ ≥10 and DP ≥10, across samples) (right y-axis, black points). **B)** Number of genomic features well covered by lrGS only across gene groups, as well as ClinVar P/LP variant positions (defined as median coverage ≥90% in lrGS and <90% in srGS). **C)** Examples of exons in disease-associated genes well covered by lrGS only, displaying read alignments in IGV for the srGS and corresponding lrGS in one individual.

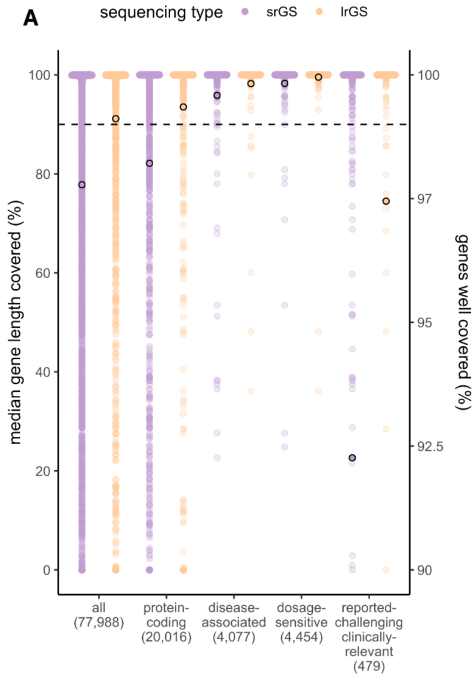


**B**

| | | N |
|---|---|---|
| genes well covered by lrGS only | all | 1,143 |
| | protein-coding | 260 |
| | disease-associated | 11 |
| | dosage-sensitive | 6 |
| | reported-challenging clinically-significant | 26 |
| genes with ≥1 exon well covered by lrGS only | protein-coding | 309 |
| | disease-associated | 20 |
| | dosage-sensitive | 8 |
| | reported-challenging clinically-significant | 35 |
| ClinVar 2-star P/LP variant positions well covered by lrGS only | | 39 |

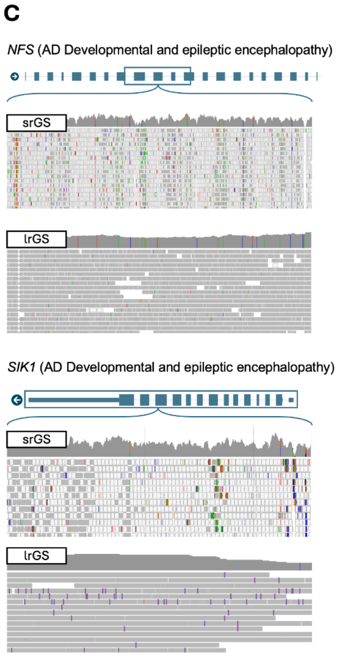

**Figure S8. Episignature concordance** for 16p11.2 deletions and CHD8 variants from Siu et al.[9] across the cohort, with affected or unaffected status indicated. The dotted line represents the threshold for a match (as previously recommended).[10] Proband RGP_2159_3 is highlighted and shows no concordance with these episignatures.

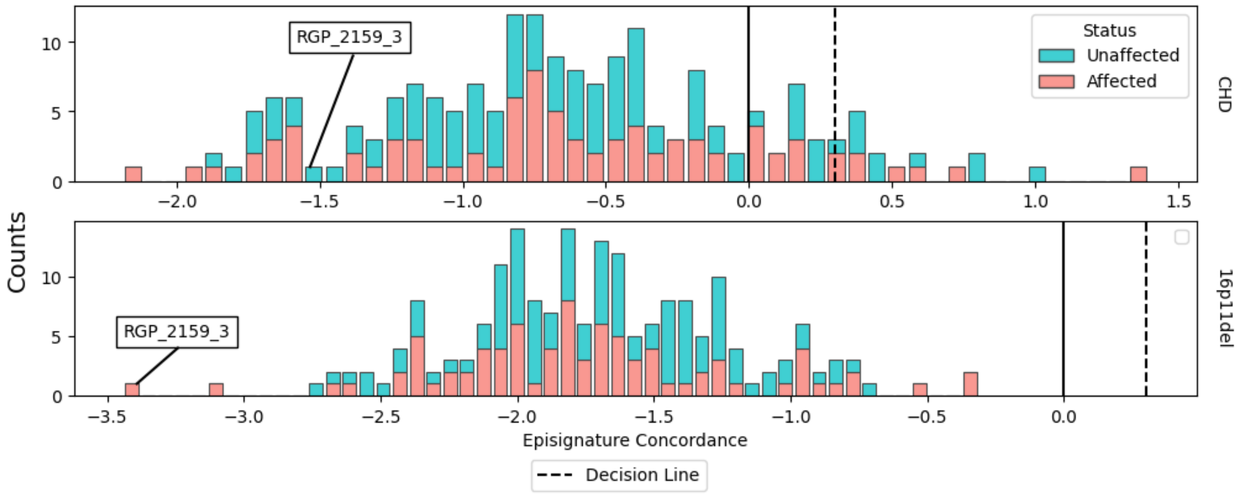

**Figure S9. Transcript classification count per sample from long-read sequencing.** Counts of transcripts identified in each sample separated by SQANTI3 classifications before **A)** and after **B)** using the SQANTI3 filters.

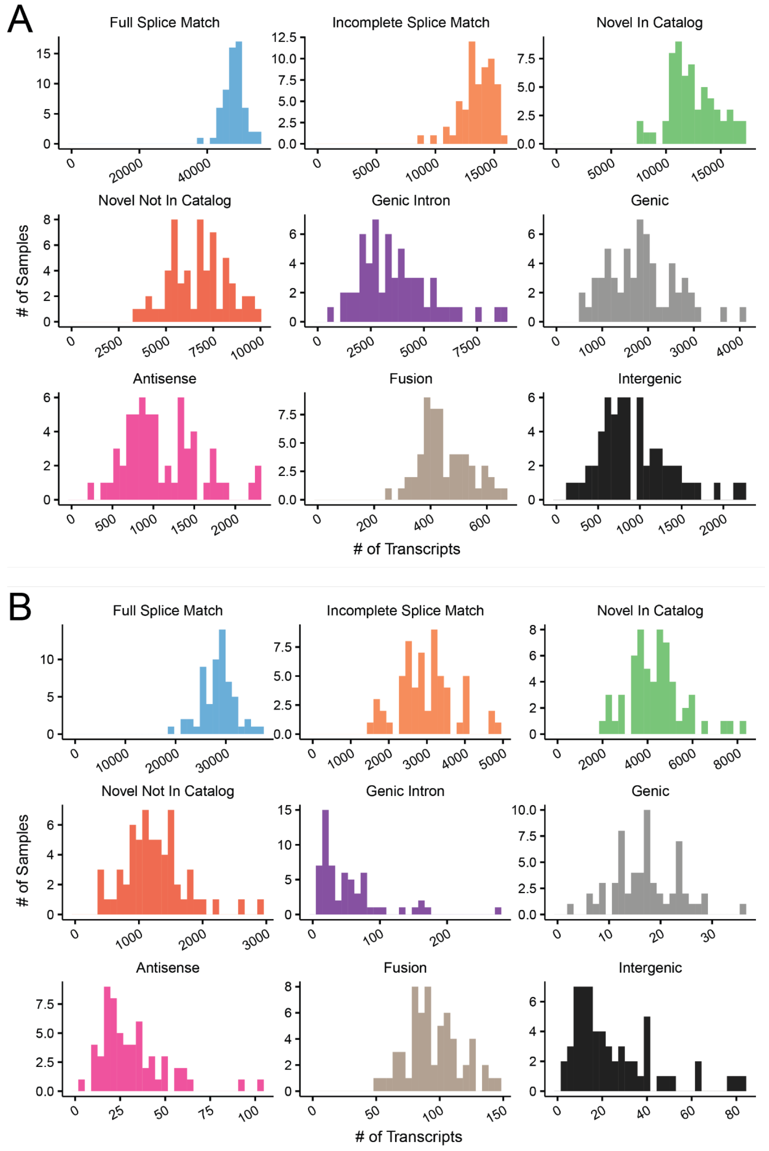
